# Genome-wide significant risk loci for mood disorders in the Old Order Amish founder population

**DOI:** 10.1101/2022.02.22.22271369

**Authors:** Elizabeth M. Humphries, Kwangmi Ahn, Rachel L. Kember, Fabiana L. Lopes, Evelina Mocci, Juan M. Peralta, Bipolar Sequencing Consortium, John Blangero, David C. Glahn, Fernando S. Goes, Peter P. Zandi, Peter Kochunov, Cristopher Van Hout, Alan R. Shuldiner, Toni I. Pollin, Braxton D. Mitchell, Maja Bucan, L. Elliot Hong, Francis J. McMahon, Seth A. Ament

## Abstract

**Background:** Genome-wide association studies (GWAS) of mood disorders in large case-control cohorts have identified numerous risk loci, yet pathophysiological mechanisms remain elusive, primarily due to the very small effects of common variants.

**Methods:** We sought to discover risk variants with larger effects by conducting a genome-wide association study of mood disorders in a founder population, the Old Order Amish (OOA, n=1,672).

**Results:** Our analysis revealed four genome-wide significant risk loci, all of which were associated with >2-fold relative risk. Quantitative behavioral and neurocognitive assessments (n=314) revealed effects of risk variants on sub-clinical depressive symptoms and information processing speed. Network analysis suggested that OOA-specific risk loci harbor novel risk-associated genes that interact with known neuropsychiatry-associated genes via gene interaction networks. Annotation of the variants at these risk loci revealed population-enriched, non-synonymous variants in two genes encoding neurodevelopmental transcription factors, *CUX1* and *CNOT1*.

**Conclusions:** Our findings provide insight into the genetic architecture of mood disorders and a substrate for mechanistic and clinical studies.

## INTRODUCTION

Mood disorders, including major depressive disorder (MDD) and bipolar disorder (BD), affect more than 300 million people worldwide (1). Identifying genetic risk factors is a promising path toward pathophysiological mechanisms and novel therapeutic targets, with genetic factors estimated to account for 60-80% (2,3) and 30-50% (4,5) of risk in BD and MDD, respectively. Genome-wide association studies (GWAS) in large case-control cohorts have revealed 64 genome-wide significant risk loci for BD and 178 for MDD, and have documented strong genetic correlations between BD and MDD (6–11). However, the effect sizes of individual risk variants are extremely small, collectively explaining at most 10 to 20% of the observed heritability (6–13). The causal mechanisms at most of these loci remain speculative, and few have been functionally characterized. Thus, the genetic causes and biological mechanisms of mood disorders remain poorly understood.

Population bottlenecks in founder populations lead to the enrichment of many functional alleles that are rare in the broader population (14–16). Some of these alleles may have larger effects on disease risk than common variants typically identified through GWAS in the broader population. The Lancaster Old Order Amish (OOA) are conservative Anabaptists who comprise a closed founder population of ∼40,000 individuals living primarily in Lancaster County, Pennsylvania (17–20). Genetic studies in this population have led to the discovery of risk variants and pathophysiological mechanisms for numerous complex and Mendelian traits (14,21–23). Genetic studies of mood disorders in the OOA were initiated in the 1970s, primarily within the Amish Study of Major Affective Disorders (ASMAD). Initial studies in this cohort identified suggestive linkage peaks, while more recent genome sequencing studies suggested polygenic effects of single-nucleotide variants and copy number variants (21,22,24–27). However, previous studies were limited by their small sample sizes (n < 400).

Here, in an expanded OOA cohort (n = 1,672), we describe the first genome-wide significant risk loci for mood disorders in this population. We provide evidence that these associations are driven by population-enriched founder alleles with large effects. We further assessed effects of these variants on quantitative behavioral and cognitive sub-phenotypes, identified convergent effects on neuropsychiatry-related gene networks, and discovered functional variants at the risk loci that are predicted to impact neurodevelopmental genes.

## METHODS AND MATERIALS

### Cohorts and genotyping

We performed whole-genome genotyping of two newly collected OOA cohorts comprised of multiply-affected pedigrees with mood disorders: the Amish Connectome Project (ACP) and the Amish Mennonite Bipolar Genetics Study (AMBiGen). We integrated these data with existing genotyping data from a third OOA mood disorders cohort, the Amish Study of Major Affective Disorders (ASMAD) (22,26) and with whole-genome sequencing (WGS) of population controls from the Amish Cohort of the Trans-Omics for Precision Medicine program (Amish TOPMed) (28,29). Table S1 provides details of the cohorts and genotyping.

### Data processing

Uniform processing, quality control, and imputation of the ACP, AMBiGen, and ASMAD genotypes was performed as previously described for ASMAD (22,26). Briefly, quality control within each cohort prior to imputation included removing SNPs missing from more than 2% of individuals, as well as those with a minor allele frequency less than 0.2% and HWE p-value less than 1 × 10^−6^ using the –geno, --maf, and –HWE commands in PLINK v1.9(30,31). Individuals missing more than 5% of SNPs or with heterozygosity greater than 3 standard deviations from the mean were removed (--missing and –het commands, respectively). Allele frequencies were checked against the Haplotype Reference Consortium and 1000 Genomes using perl commands provided by the Wellcome Sanger Institute (32). Imputation was performed on the Michigan Imputation Server(33) using the TOPMed Freeze5 reference panel, which includes WGS from the Amish TOPMed cohort among ∼65,000 genomes. We used the GRC38/hg38 build with a European population, no r-square filtering, and Eagle v2.4 phasing, using the quality control and imputation mode. We removed all non-polymorphic sites from both the imputed and directly sequenced genomes, then renamed all remaining sites by chromosome, position, reference allele, and alternate allele using bcftools annotate(34). Finally, polymorphic-subsetted datasets were merged (PLINK v1.9(30,31)--merge-list). We filtered out all imputed SNPs with an imputation r^2^ < 0.6.

### Assessment of population structure

We calculated principal components for the genomes using the –pca command in PLINK v1.9 (30,31), after removing SNPs missing from more than 5% of the entire sample and with a minor allele frequency less than 1% (--geno and –maf). This analysis was performed using the imputed genomes for the ACP, AMBiGen, and ASMAD cohorts and the WGS from the TOPMed cohort as there were only 598 polymorphic SNPs in common among the four genotyping panels. PC1 separated the Lancaster OOA from various non-OOA populations collected in the ACP and AMBiGen studies. We removed all individuals that did not belong to the Lancaster OOA population, then re-calculated PCA. Lancaster OOA-specific PCs were used as covariates in the GWA analysis.

### Assessment of sample overlap

We calculated identical-by-descent (IBD) allele sharing statistics on Lancaster OOA samples with the PLINK v1.9 –genome command (30,31). We used the proportion of IBD values to identify individuals who were enrolled in more than one cohort. Samples with a proportion value > 0.8 were assumed to be from the same individuals, and duplicate samples were removed. For each individual, the most recent, most-deeply-phenotyped sample was retained (ACP > AMBiGen > ASMAD > TOPMed; Table S1).

### Assessment of genotyping and imputation accuracy

Accuracy of imputed genotypes was confirmed through comparison to WGS performed for a subset of the individuals in each cohort. These validation datasets included Illumina WGS (∼30x average coverage) for 214 of the individuals in the ACP cohort obtained as part of the Whole-Genome Sequencing of Psychiatric Disorders consortium (David Glahn and John Blangero, PIs); Complete Genomics WGS for 80 participants in ASMAD (22,26); and Illumina WGS (∼30x) for 93 individuals enrolled in both the Amish TOPMed cohort and one of the mood disorders cohorts. Details of sequencing, genome alignment, and variant calling for the ASMAD and TOPMed WGS have been described (22,27–29). For the ACP WGS, whole-genome sequencing was performed on an Illumina HiSeq-X at the Broad Institute of MIT and Harvard. Reads were aligned to the hg38 reference genome, and variant calling was performed jointly across all samples from this cohort using freebayes (35) (v1.3.1) with the following parameters: use-best-n-alleles 3, min-alternate-count 5, - -min-alternate-fraction 0.2, --min-coverage 10, and --limit-coverage 500. We note that while the sequencing and genotyping-based variant calls for ACP and ASMAD are fully independent, the TOPMed WGS are not fully independent due to the inclusion of these 93 individuals in the imputation panel. Treating the ACP and TOPMed WGS as a gold standard, we calculated the precision and recall for non-reference genotype calls across all alleles. In addition, in all cohorts we specifically verified the genotypes for the four lead SNPs at genome-wide significant risk loci: rs192622352, rs569742752, rs117752843, and rs7185072.

### Affection status models

The primary phenotype was diagnosis with a bipolar spectrum disorder, including individuals with primary diagnoses of Bipolar Disorder Type I (n=86), Bipolar Disorder Type II (n=17), Bipolar Disorder Not Otherwise Specified (n=10), or Recurrent Major Depressive Disorder (n=73). We did not include Single Episode Major Depressive Disorder in this phenotype because the heritability of this disorder is much lower than the heritability of Recurrent Major Depressive Disorder (36). Individuals from the AMBiGen, ASMAD, and ACP cohorts (the cohorts ascertained on mood disorders) were coded as unaffected if they had no Axis I or Axis II diagnosis (n=449). All individuals from the TOPMed general population cohort were coded as unaffected (n=938). In the primary analysis, individuals with other Axis I or Axis II diagnoses were coded as unknown, and we considered these diagnoses in alternative affection status models, as follows. The cohort included ten individuals with a primary diagnosis of schizophrenia, all of whom were first- or second-degree relatives of mood disorder cases. While SCZ is not classically identified as a mood disorder, there is substantial genetic, clinical, and brain pathophysiological overlaps between SCZ and mood disorders, especially bipolar disorders (37,38). Therefore, we studied an alternative affection status model in which individuals with SCZ were coded as affected. In addition, we tested models in which Single-Episode Major Depressive Disorder and Persistent Depressive Disorder were coded as affected. Individuals from these cohorts with a psychiatric diagnosis other than the diagnoses above or who did not undergo a psychiatric evaluation were always coded as unknown (n=62). We also considered a model in which only individuals with recurrent MDD were coded as affected, as well as a model in which only individuals with a BD diagnosis were coded as affected.

### Genome-wide association analysis

We tested associations of genotyped and imputed variants with mood disorders, as defined above, in our sample of 1,672 Lancaster OOA individuals using a mixed-effect linear regression model implemented with EMMAX (39,40). Covariates included an empirical kinship matrix and twenty principal components, which account for family structure and more distant relatedness, respectively.

### LD clumping

We identified linkage disequilibrium (LD)-independent lead SNPs and sets of genetically correlated SNPs in the Old Order Amish using the –clump command in PLINK v1.9 (30,31), setting the significance threshold for lead SNPs to 1×10^−5^ and the secondary significance threshold for clumped SNPs to 0.05. We set the LD threshold to 0.6 and the physical distance threshold to 1000 kb. We also allowed for non-index SNPs to appear in multiple loci. After we generated the list of loci, we identified SNPs and indels in LD with each of the lead SNPs. For this purpose, we utilized WGS from OOA participants in the ACP study, so as to include unimputed, population-specific variants. We used D’ as our primary measure of linkage disequilibrium, enabling us to identify linked variants that differ in allele frequency from the lead SNPs (e.g., variants on sub-haplotypes). This analysis was performed with a call to PLINK v1.9 with the following flags: --r2 dprime with-freqs --ld-snp chr7:103511937:C:T --ld-window 100000 --ld-window-kb 20000 --ld-window-r2 0.05. We note that in all of these analyses of LD, the physical distance thresholds were set to larger values than is typical of GWAS in the broader population due to the longer haplotypes in this founder population.

### Pseudo-replication

We performed pseudo-replication analyses using a leave-one-out strategy to verify that the results are not dependent on samples from a single cohort. We removed one cohort at a time (ACP, AMBiGen, ASMAD, or TOPMed) from the sample and reran EMMAX(39). We recalculated PCA coordinates for each pseudo-replication dataset and used the first 20 recalculated coordinates as covariates in the model. We also re-ran the analysis on the ACP and ASMAD cohorts independently, again using 20 recalculated PCs as covariates.

### Annotation of loci and variants

We assessed overlap of risk loci identified in the OOA with loci from published large-scale neuropsychiatric GWAS. We used the BEDtools v2.27.1(41) intersect command to calculate the overlap between the risk-associated loci (defined as SNPs with r^2^ > 0.6 to one of the 4 lead SNPs) and risk-associated SNPs identified in previous GWAS of mood disorders and related neuropsychiatric traits: MDD(7), BD(8,9), SCZ(42), and educational attainment(43). We used the authors’ definitions of risk loci for MDD, BD, and SCZ. For the educational attainment dataset, bounds of risk loci were not described in the original publication, so we set bounds 250 kb upstream and downstream of the lead SNPs. We also tested whether the lead SNPs identified in the Lancaster OOA sample were in LD with risk-associated SNPs identified in previous neuropsychiatric GWAS using the PLINK’s –ld command and recorded the r^2^ value.

We further annotated proximal candidate genes at risk loci using gene sets related to autism spectrum disorders (ASD), BD, and SCZ. Within the bounds of each risk locus, we annotated differentially expressed genes in the prefrontal cortex of ASD, BD, and SCZ cases vs. controls from PsychENCODE(44). We also annotated genes from exome sequencing studies, including genes with a gene burden p-value < 2.5 × 10^−6^ from SCHEMA(45) (SCZ-associated genes), and ASD-associated genes from Satterstrom *et al*.(46) and SFARI Gene(47).

We annotated the variants at each locus using the Ensembl Variant Effects Predictor (VEP, release 105, accessed online, January 31, 2022) with the following parameters: assembly GRCh38.p13, Ensembl/GENCODE transcripts, 1000 Genomes global and continental allele frequencies, gnomAD exomes allele frequencies, and including CADD scores.

### Polygenic risk score (PRS) analysis

We calculated polygenic risk scores for each OOA individual, using results from large-scale GWAS of BD(8,9), SCZ(42), and MDD(7) using PRSice-2(48). SNPs with a minor allele frequency less than 0.05, Hardy-Weinberg equilibrium p-value less than 1 × 10^−6^, or missing in more than 10% of individuals were removed from the dataset before analysis, as well as individuals missing more than 10% of SNPs. Any SNPs removed due to these filters were removed from the entire analysis, so every individual had the same number of SNPs used to calculate their score. We calculated PRS using the full Lancaster OOA dataset, as well as for each mood disorder cohort (ACP, AMBiGen, and ASMAD) dataset separately and used the PRS calculated at the program-estimated best threshold (BD: 0.02825; SCZ: 0.0392; MDD: 0.00015). We constructed logistic regression models with the glm() function in the R stats package(49) to test for additive and non-additive effects of these PRS and of OOA-specific risk alleles on affection status, including 20 PCs as covariates. We compared models with and without a PRS x lead SNP interaction term to evaluate non-additive effects.

### Effects of risk variants on quantitative behavioral and neurocognitive phenotypes

We tested for the associations of the lead SNPs at genome-wide significant risk loci identified by the GWA analysis with quantitative behavioral and cognitive traits in 314 OOA participants in the ACP study, including 84 individuals affected with a mood disorder. We studied self-reports of current depression symptoms from the Beck Depression Inventory (50), lifetime depression symptoms from the Maryland Trait and State Depression scale (51), and lifetime history of bipolarity from the Bipolar Spectrum Diagnostic Scale (52). We also used scores from several cognitive tasks, including digit sequencing (verbal working memory), digit symbol coding (processing speed, visuospatial memory), spatial span (visuospatial working memory), and the Wechsler Abbreviated Scale of Intelligence(53) (WASI) matrix reasoning and vocabulary subtests (IQ and cognitive ability). We assessed normality, as well as associations of each trait with age and sex. The scores from Beck Depression Inventory, Bipolar Spectrum Diagnostic Scale, and spatial span were transformed using a square root transformation to improve normality. The other five traits displayed non-linear associations with age. For those traits, we applied a loess regression model (using the loess function in R v. 3.6.2(49), span = 0.5) and performed genetic association tests on residuals. Covariates in the EMMAX model for these three traits included sex, age, and an empirically constructed kinship matrix. The heritability of each trait was calculated using SOLAR-Eclipse(54). We constructed mixed-effect linear regression models for each genotype-phenotype pair using EMMAX (39,40).

### Gene set enrichment analysis

Gene-based p-values were computed from GWAS summary statistics using MAGMA (55). SNPs were annotated to ENSEMBL genes, including a 10 kb window up- and downstream of each gene’s genomic coordinates. Gene p-values were computed using the lowest SNP p-value as the test statistic (snp-wise=top,1), and gene-gene correlations were computed using our imputed OOA genotype matrix. MAGMA gene set enrichment analyses were performed with default parameters. We studied 21 gene sets with prior evidence for association with neuropsychiatry, as described previously (56,57). Briefly, these gene sets were derived from the following sources: genes identified through GWAS of MDD(7), BD(8), SCZ(58), and neuroticism(59); genes identified through genetic association studies of rare variants, including exome and genome sequencing studies of SCZ(45), autism spectrum disorders (ASD) (46), or other developmental disorders(60), as well as genes intolerant to loss-of-function mutations(61); genes that are differentially expressed in the prefrontal cortex of individuals with BD, SCZ, or ASD (44); genes that have been identified as targets of the RNA binding proteins FMRP, RBFOX2, RBFOX1/3, and CELF4, of the chromatin remodeling genes CHD8, and of the microRNA miR-137 (57,62); genes localized to synapses from SynaptomeDB (63).

### Gene interaction networks

Human protein-protein interactions were downloaded from the STRING database (64) (https://stringdb-static.org/download/protein.links.detailed.v11.5/9606.protein.links.detailed.v11.5.txt.gz). We defined OOA risk genes as those with a MAGMA gene p-value < 0.01. We used the same 21 neuropsychiatry-related gene sets as in MAGMA gene set enrichment analysis. To assess interactions between established gene sets and OOA risk genes, we counted the number of protein-protein interactions that directly link OOA risk genes to genes in each of the 21 established neuropsychiatry gene sets. We tested whether the number of interactions was greater than expected by chance by two approaches. First, we computed Fisher’s exact tests. Second, we repeatedly permuted the edges of the network, holding each node’s degree constant, and compared the number of OOA-known edges in observed vs. permuted data. Edge permutations were used to confirm results from Fisher’s exact test (n=100 permutations). Odds ratios and p-values from Fisher’s exact test are reported in the manuscript, as they provide a more precise measure of the likelihood.

To prioritize specific OOA risk genes, we ranked them by their centrality within a gene interaction network centered on known neuropsychiatry risk genes. We defined a set of 684 core neuropsychiatry genes with evidence from at least three independent approaches from our 21 gene sets, as follows: Genes implicated by studies of rare variants, defined as the union of genes associated with disease in exome sequencing studies of SCZ and ASD. Genes implicated by gene expression profiling were defined as the union of genes that were significantly down- or up-regulated in prefrontal cortex of BD, SCZ, or ASD cases using data from psychENCODE. Genes implicated by gene network analyses were defined as the union of genes that are targets of CELF4, FMRP, RBFOX1/3, RBFOX2, CHD8, and miR-137. Synaptic genes were defined from the SynaptomeDB database. We excluded genes derived from GWAS, as these genes are potentially non-independent from association signals in our OOA dataset. We extracted all protein-protein interactions from the STRING database for which at least one node was one of these 684 genes. In practice, the large number of interactions in the STRING database means that nearly all genes are represented in this network, but only the subset of their interactions that involve neuropsychiatry-related genes. We used the eigen_centrality() function from the igraph R package (65) to calculate the centrality of each node, including OOA risk genes that have not previously been implicated in neuropsychiatric disorders. We computed ranks for the OOA risk genes, separately, based on eigen-centrality, as well as based on their MAGMA p-values. The final ranking is the median rank from these two metrics. We tested for functional enrichments within the top 250 genes from this analysis using DAVID (66).

### Expression of CUX1 and CNOT1 in the human brain

We evaluated the expression of *CUX1* and *CNOT1* using RNA-seq of developing and adult brain regions (67), downloaded from the BrainSpan website (https://www.brainspan.org/static/download.html). Analyses of *CNOT1* expression utilized normalized counts summarized to Gencode v10 gene models. Analyses of *CUX1* expression utilized normalized counts summarized ton exons. We studied the expression of the following *CUX1* exons (hg19 coordinates). chr7:101921219-101921336, exon 17 of ENST00000425244.6, which contains rs118010189 and is expressed only in splice forms that encode CASP, and chr7:101891691-101901513, which spans the final exon and 3’ untranslated region of *CUX1* transcription factor-encoding transcripts.

### Data availability

Genotypic and phenotypic data from the Amish TOPMed study and from AMBiGen are available through the National Institute of Health Database of Genotypes and Phenotypes (phs000956.v1.p1, phs000899.v1.p1). Genotypic and phenotypic data from ACP are available through the NIMH Data Archive (Study #2902). Genotypic and phenotypic data from ASMAD are available through the Coriell Institute for Medical Research.

## RESULTS

### Genome-wide association study of mood disorders in the Old Order Amish founder population

We generated whole-genome genotyping data from two newly collected Anabaptist cohorts with mood disorders -- the Amish Connectome Project (ACP) and the Amish and Mennonite Bipolar Genetics study (AMBiGen) – and we integrated these with existing data from two additional cohorts -- ASMAD and the Trans-Omics for Precision Medicine cohort (TOPMed). Following uniform quality control and imputation, we studied 6.6 million polymorphic single-nucleotide polymorphisms (SNPs) in 1,672 OOA individuals from this combined cohort, of whom 196 were affected with a major mood disorder (BD, recurrent MDD, or schizoaffective disorder; Tables S1-S2). Power analyses(68) suggest that this cohort is well-powered to detect population-enriched risk alleles with moderate to large effects, equivalent to those discovered in the OOA for non-psychiatric traits(14,23,69). Principal component analysis (PCA) indicated that these OOA individuals form a discrete population compared to other Anabaptist groups in our sample (Fig. S1A), with minimal stratification by study or genotyping platform (Fig. S1B). Whole-genome sequencing (WGS; n=214 from ACP and n=87 from TOPMed) confirmed >99.9% precision for imputed non-reference genotype calls, with >99.8% recall (Table S3).

We conducted a genome-wide association study (GWAS) of mood disorder affection status in this OOA cohort using a linear mixed model implemented with EMMAX (39). Twenty-five SNPs were associated with affection status at a conventional genome-wide significance threshold, *P* < 5 × 10^−8^ (Fig. 1A). The quantile-quantile plot of observed vs. expected p-values revealed no genomic inflation, as well as an excess of p-values less than ∼1e-4, consistent with polygenicity (λ _GC_ = 0.8; Fig. S2A). Linkage-disequilibrium (LD)-based clumping supported four genome-wide significant risk loci at cytobands 3q28/29, 5q13, 7q22, and 16q21 (Fig. S2B-E; Table S4). Each of the four lead SNPs was associated with >2-fold relative risk. Consistent with founder effects, the lead SNPs or other SNPs in LD with them (D’ > 0.9) at all four loci were uncommon, OOA-enriched SNPs on extended haplotypes(15). Carriers from 3-10 families contributed to each allele’s association with mood disorders (Figs. 1B, S3A-B).

**Figure 1.**
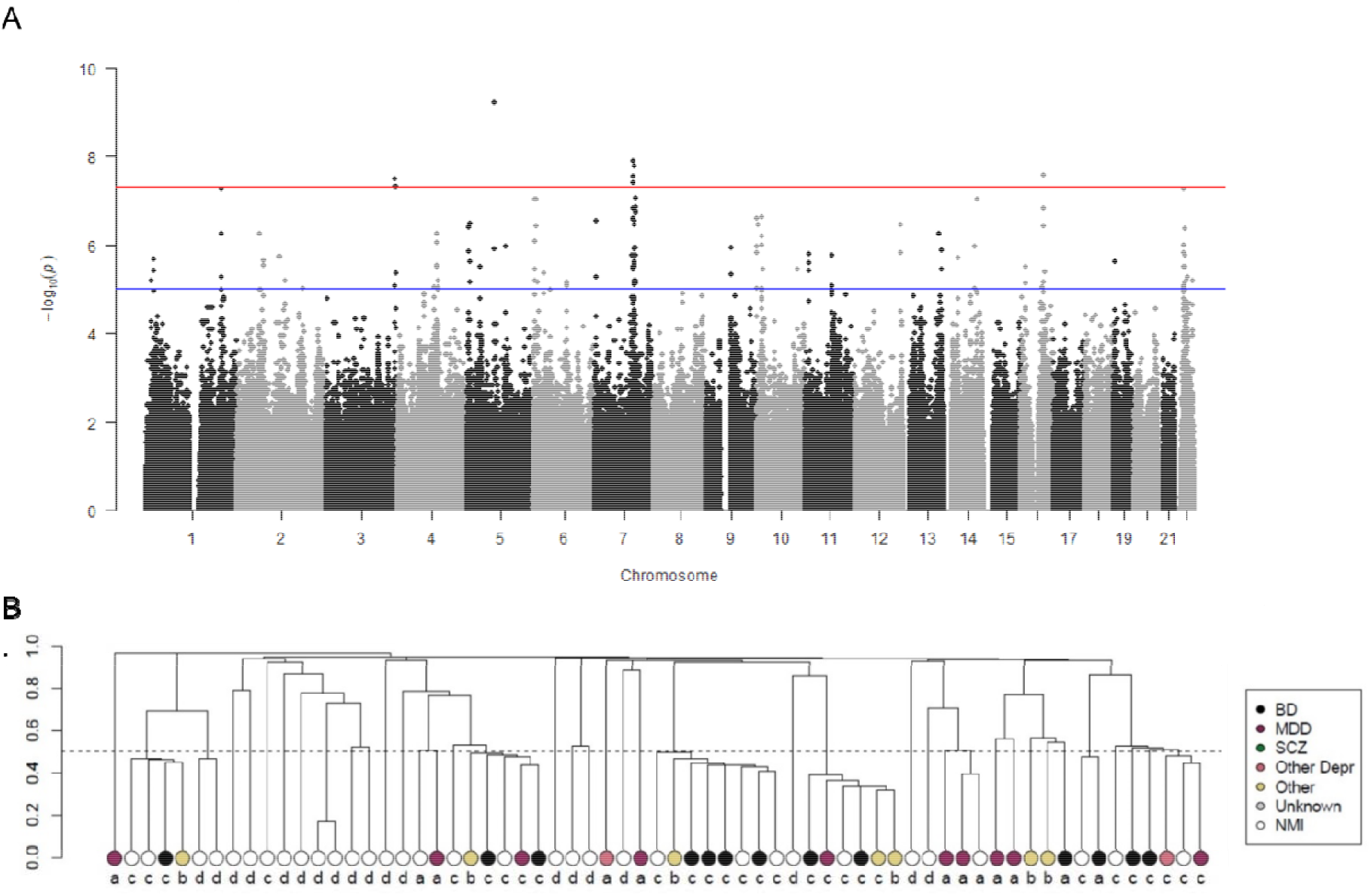
Genome-wide association study of mood disorders in the Old Order Amish founder population (n=1,672). A. Manhattan plot: -log_10_(p-values) for associations of 6.6 million single-nucleotide polymorphisms (SNPs) with mood disorders. B. Empirical kinship among carriers of the lead SNP at the 7q22 risk locus. Y-axis indicates the coefficient of relatedness, with the dotted line at 0.5 indicating first-degree relatives. a=ACP, b=AMBiGen, c=ASMAD, d=TOPMed.

Several analyses support the robustness and reproducibility of these associations. First, we confirmed 100% accuracy for reference and non-reference genotype calls for all four lead SNPs by comparing the imputed genome to WGS (n=214 from ACP, n=80 from ASMAD (22,26,27), and n=87 from TOPMed). These results indicate that the associations are not an artifact of biases in genotyping or imputation.

Second, we performed pseudo-replication analyses within subsets of our OOA sample using a leave-one-cohort-out approach, in which the GWAS was conducted using the integrated data from all but one cohort. All four lead SNPs remained nominally significant (*P* < 0.05; Table S5). The lead SNPs at 5q13, 7q22, and 16q21 were also nominally significant in GWAS of the ASMAD and ACP cohorts singly. Carriers of the 3q28/29 lead SNP were identified primarily in ACP, with a single affected carrier in the ASMAD cohort. These results indicate that the associations are reproducible in multiple OOA cohorts.

Third, we performed secondary analyses using alternative affection status models (Table S6). All four lead SNPs remained either significant (*P* < 5 × 10^−8^) or suggestive (*P* < 5 × 10^−6^) when we broadened the affection status model to include Persistent Depressive Disorder and Single Episode MDD, rather than removing these individuals from the analysis. Similar results were obtained when we treated ten participants with SCZ as affected (rather than excluding them from the analysis). We also found suggestive associations for all four lead SNPs when we considered only recurrent MDD cases to be affected and when we considered only BD cases to be affected. These results suggest that the associations are robust to affection status model and that the loci influence risk for multiple mood disorders.

Fourth, we performed analyses to test whether the risk loci identified in the OOA overlap known risk loci for mood disorders and related traits in the broader population (7)(8,9) (42) (43) (Table S7). The 5q13 risk locus overlaps previously reported risk loci for BD and educational attainment. The 16q21 risk locus overlaps a previously reported risk locus for educational attainment. The 7q22 risk locus overlaps previously reported risk loci for MDD, BD, SCZ, and educational attainment. Though the 3q28/29 risk locus has not previously been identified in large-scale GWAS of BD, MDD, SCZ, or educational attainment, 3q29 microdeletions are associated with increased risk for BD and SCZ (70,71). By contrast, none of the lead SNPs or genetically correlated SNPs identified in the OOA had significant p-values in previous GWAS of mood disorders. This Is expected, as causal alleles whose true effect sizes are large are expected to be selected against and rare in the broader population, and the lead SNPs at three of the four risk loci are uncommon even in the OOA. Taken together, these results suggest that the loci we identified in the OOA correspond to novel risk haplotypes, potentially with large effects, at known risk loci for mood disorders and related traits.

### Evaluation of genotype-phenotype relationships with polygenic risk scores and deep phenotyping

The discovery of risk alleles with substantial effects provides opportunities for deeper exploration of genotype-phenotype relationships. First, we evaluated the relationship between OOA-specific risk alleles and polygenic risk from common variants. Consistent with previous studies in the ASMAD cohort (22,26), a polygenic risk score (PRS) for BD, derived from GWAS in the broader population (8,9), explained a small but significant proportion of risk for mood disorders in our cohort (2.4% of risk, *P =* 8.3×10^−6^; Fig. S4), corresponding to a 2.6-fold relative risk in the top vs. bottom quartile. Individuals with mood disorders had significantly higher PRS than their unaffected family members (*P* = 9.4 × 10^−6^). Similar results were obtained using PRS for SCZ and MDD (Fig. S4). We tested for additive and non-additive effects of bipolar disorder PRS and OOA-specific risk alleles using multivariate logistic regression models. We found significant main effects of PRS and of each OOA-specific risk allele (*P* < 0.05), but interactions between PRS and OOA-specific alleles were not significant. These results suggest that OOA-specific risk alleles and common risk alleles have additive, independent effects on risk for mood disorders in this cohort.

Next, we tested whether OOA-specific risk alleles for mood disorders also have quantitative effects on the classic behavioral symptoms of mood disorders, which we assessed via three rating scales (n up to 314 ACP participants): the Beck Depression Inventory (50) (BDI), which measures depression symptoms in the two weeks prior to testing; the Maryland Depression Trait Scale (51) (MDTS), which assesses lifetime depression symptoms; and the modified Bipolar Spectrum Diagnostic Scale (52) (BSDS), which measures the polarity of the depressive and manic symptoms. We found significant broad-sense heritability in this sample for all three scales (*h*^*2*^ = 0.25-0.41, *P* < 0.003; Table S8) and confirmed that participants with mood disorder diagnoses had higher scores (Table S8). The lead SNPs at the 3q28/29, 7q22, and 16q21 risk loci were all associated with higher scores on the MDTS (Fig. 3, Table S9). Also, the 7q22 lead SNP was associated with a higher BSDS score. However, none of the lead SNPs were significantly associated with BDI, suggesting these loci more strongly influence lifetime history than current symptoms. These results suggest that the OOA-specific risk alleles identified in our analysis impact the core behavioral symptoms of MDD and BD.

**Figure 2.**
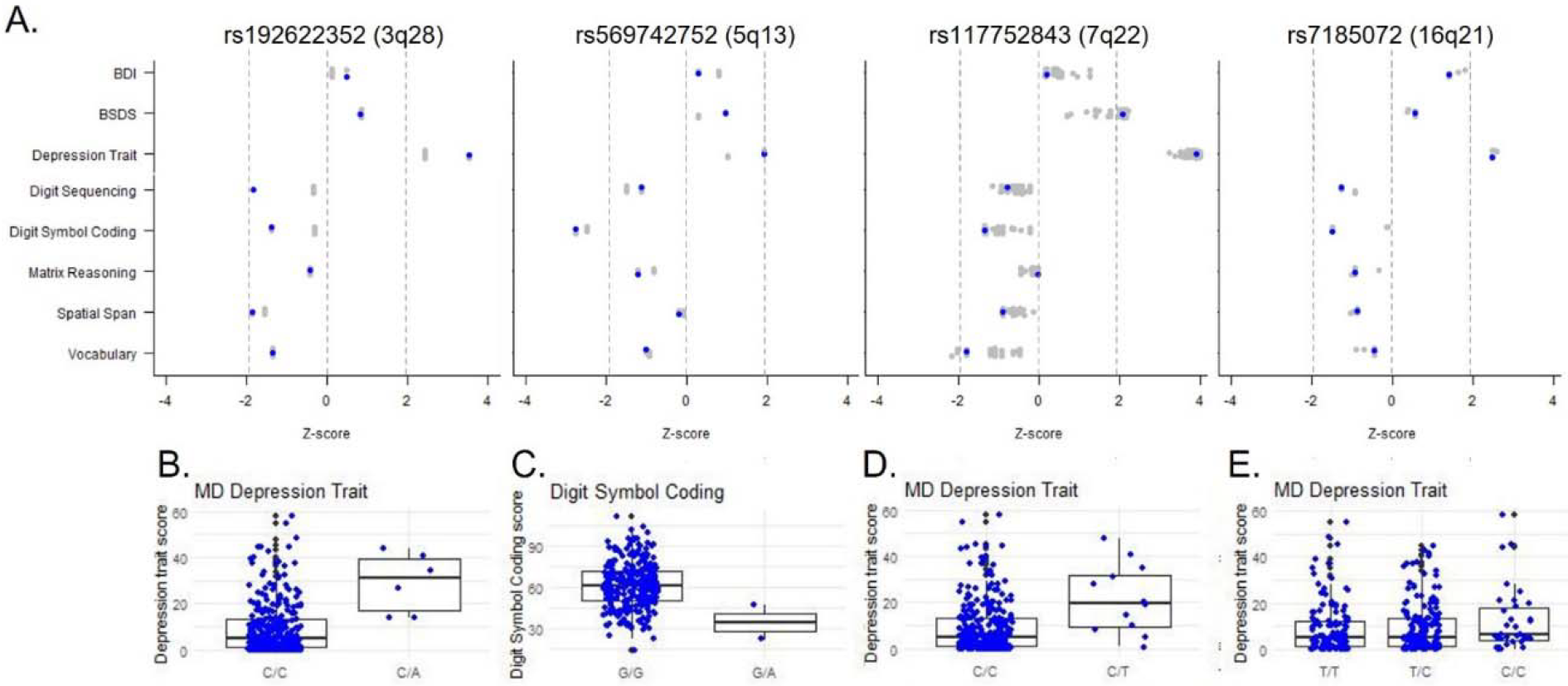
Associations of OOA-specific risk alleles for mood disorders with depressive symptoms and cognitive performance. A. Direction and significance (z-score) of each SNP’s associations with behavioral and cognitive traits. Blue dots indicate lead SNPs, and gray dots indicates other SNPs in LD with the lead SNP at each locus. BDI = Beck Depression Inventory; BSDS = bipolar spectrum diagnostic scale; depression trait = Maryland Depression Trait Scale. B-E. Plots of the most strongly associated trait for each lead SNP.

**Figure 3.**
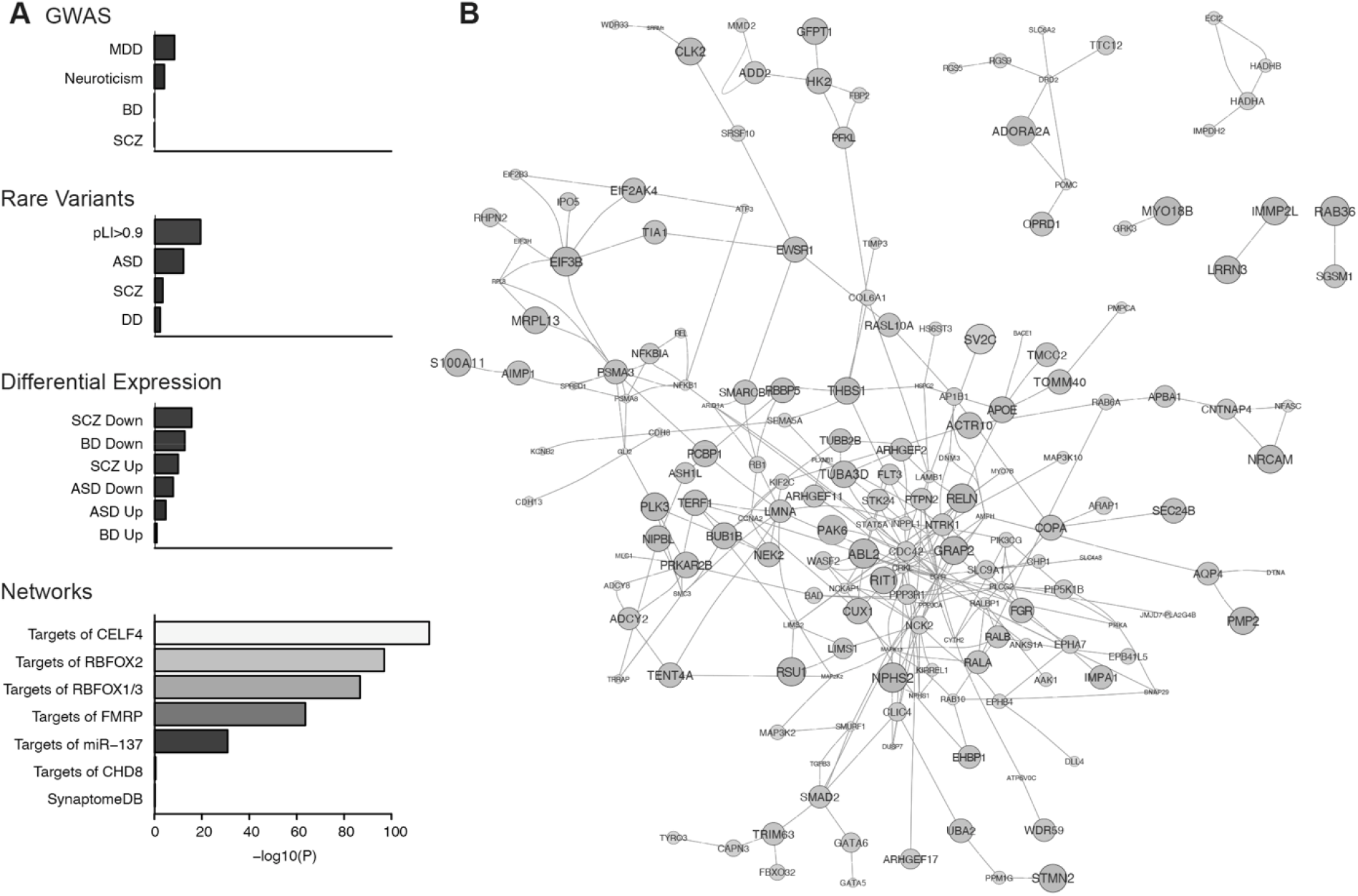
Genes at OOA-specific risk loci for mood disorders interact with neuropsychiatry-related gene networks. A. Putative OOA risk genes (820 genes; MAGMA, *P* < 0.01) had an elevated rate of protein-protein interactions with gene sets derived from GWAS, rare variant studies, differential gene expression, and network analyses of psychiatric disorders. B. Protein-protein interactions among the top 250 genes at OOA risk loci prioritized by their centrality in a gene network centered on known neuropsychiatry-related genes and strength of their statistical association with mood disorders in the OOA. Node size and color correspond to MAGMA p-values.

**Table 1.**
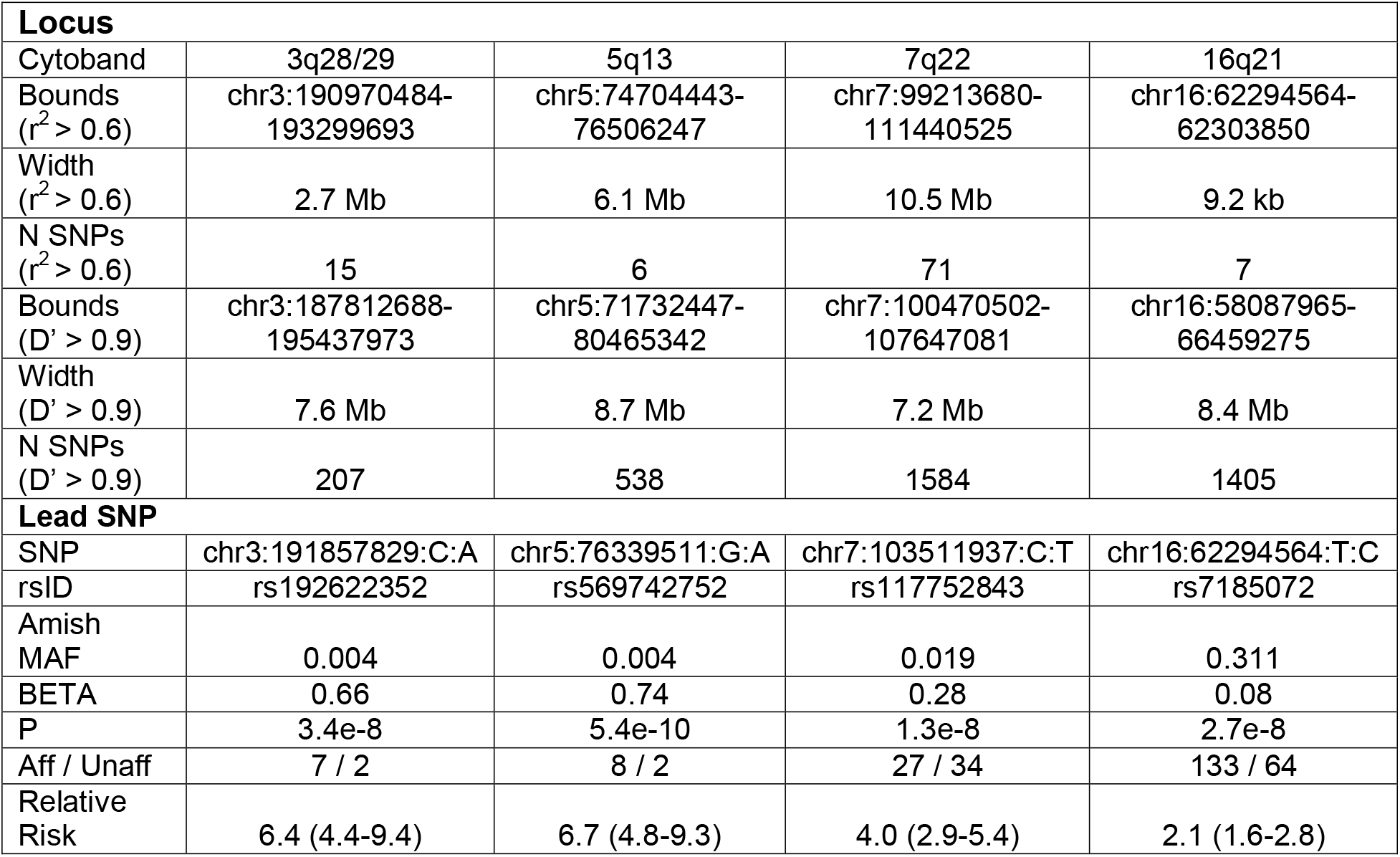
Summary of risk loci.

| **Locus** |  |  |  |  |
| --- | --- | --- | --- | --- |
| Cytoband | 3q28/29 | 5q13 | 7q22 | 16q21 |
| Bounds<br>($r^2 > 0.6$) | chr3:190970484-193299693 | chr5:74704443-76506247 | chr7:99213680-111440525 | chr16:62294564-62303850 |
| Width<br>($r^2 > 0.6$) | 2.7 Mb | 6.1 Mb | 10.5 Mb | 9.2 kb |
| N SNPs<br>($r^2 > 0.6$) | 15 | 6 | 71 | 7 |
| Bounds<br>($D' > 0.9$) | chr3:187812688-195437973 | chr5:71732447-80465342 | chr7:100470502-107647081 | chr16:58087965-66459275 |
| Width<br>($D' > 0.9$) | 7.6 Mb | 8.7 Mb | 7.2 Mb | 8.4 Mb |
| N SNPs<br>($D' > 0.9$) | 207 | 538 | 1584 | 1405 |
| **Lead SNP** |  |  |  |  |
| SNP | chr3:191857829:C:A | chr5:76339511:G:A | chr7:103511937:C:T | chr16:62294564:T:C |
| rsID | rs192622352 | rs569742752 | rs117752843 | rs7185072 |
| Amish<br>MAF | 0.004 | 0.004 | 0.019 | 0.311 |
| BETA | 0.66 | 0.74 | 0.28 | 0.08 |
| P | 3.4e-8 | 5.4e-10 | 1.3e-8 | 2.7e-8 |
| Aff / Unaff | 7 / 2 | 8 / 2 | 27 / 34 | 133 / 64 |
| Relative<br>Risk | 6.4 (4.4-9.4) | 6.7 (4.8-9.3) | 4.0 (2.9-5.4) | 2.1 (1.6-2.8) |

Cognitive deficits are observed in a subset of individuals with BD and MDD (53). We assessed effects on cognition via five tasks that measure cognitive dimensions previously implicated in mood disorders: Digit Sequencing, which primarily measures verbal working memory; Spatial Span, which measures visuospatial working memory; Digit Symbol Coding, which primarily measures processing speed; Matrix Reasoning, which measures fluid intelligence, spatial ability, and perceptual organization; and Vocabulary, which measures semantic knowledge and verbal comprehension. We confirmed significant broad-sense heritability for all five tests (Table S8). Mood disorder diagnoses were associated with lower scores, especially for digit sequencing and digit symbol coding (Table S9). Despite low n, we detected an association of the lead SNP at the 5q13.3 locus, rs569742752, with decreased performance on the digit symbol coding task (n=2 carriers and 299 non-carriers, β = -28.3, *P* = 0.006). The lead SNPs at the other loci were not significantly associated with cognitive performance. Therefore, cognitive deficits are present in a minority of carriers with these risk variants.

### Gene networks associated with mood disorders in the OOA

We applied a network analysis approach to gain insight into the biological characteristics of the genes located at risk loci. As a starting point, we computed gene-based p-values from our GWAS summary statistics with MAGMA (55). This analysis revealed three exome-wide significant genes: *ATP13A5* at 3q29 (*P* = 1.6e-7), *SV2C* at 5q13 (*P* = 2.8e-7), and *MB21D2* at 3q28 (*P* = 6.5e-7), and 820 genes reaching a nominal level of significance, *P* < 0.01 (Table S10). *ATP13A5* encodes ATPase 13A5, which is highly expressed in brain pericytes and is involved in the transport of diverse cargo across cellular membranes (72). *SV2C* encodes Synaptic Vesicle Glycoprotein 2C, which is expressed specifically on the vesicles of dopaminergic neurons and contributes to dopamine release (73). *MB21D2* encodes Mab-21 Domain Containing 2.

Next, we asked whether these OOA risk genes overlap genes and gene networks previously implicated in neuropsychiatric disorders, using 21 gene sets derived from psychiatric GWAS, exome sequencing, post-mortem prefrontal cortex gene expression, and analyses of disease-associated gene networks (56,57). We tested both for direct overlap of OOA risk genes with these gene sets, as well as network overlap, in which OOA risk genes interact with established neuropsychiatry genes via protein-protein interactions (Methods). OOA risk loci contained numerous established neuropsychiatry genes. For instance, 44 genes within the bounds of the 7q22 risk locus had a prior annotation to a psychiatric disorder, including the well-established autism spectrum disorder risk genes *RELN, KMT2E*. and *CUX1*(74). However, gene-set enrichment analysis with MAGMA indicated that, overall, risk genes identified in our study were not over-represented for established neuropsychiatry gene sets (*P* > 0.05).

In contrast, we found strong evidence that OOA risk genes interact with established neuropsychiatry genes via protein-protein interactions. Specifically, we examined protein-protein interactions in the STRING database that link OOA risk genes to genes from each of the established neuropsychiatry gene sets. 15 of the 21 neuropsychiatry gene sets showed at least a nominally significant over-representation for network edges (hypergeometric test: *P* < 0.001; Fig. 4A; Table S11). The most strongly over-represented gene sets included target genes of the neuronal RNA-binding proteins CELF4 (174,778 interactions, odds ratio [OR] = 1.06, *P* = 1.0e-116) and RBFOX1 (235,084 interactions, OR = 1.05, *P* = 1.8e-87), genes down-regulated in prefrontal cortex from bipolar disorder cases (17,684 interactions, OR = 1.06, *P* = 1.8e-13), and autism spectrum disorder risk genes from exome sequencing studies (11,242 interactions, OR = 1.07, *P* = 7.8e-13). Permutations of network edges confirmed significant over-representation for each of these gene sets.

We prioritized specific OOA risk genes based on the extent of their interaction with this shared neuropsychiatry gene network. The top 250 genes, selected from among the 820 OOA risk genes with GWAS p-values < 0.01 and ranked by eigencentrality, are shown in Fig. 4B. These genes were enriched for 13 Gene Ontology functional categories (FDR < 0.01; Table S12), including genes localized to dendrites (18 genes, *P* = 5.5e-6) and genes involved in signal transduction (39 genes, *P* = 3.1e-6) and focal adhesion (18 genes, *P* = 2.5e-5). These results suggest that OOA risk loci harbor novel risk genes within a polygenic gene network that is shared with neuropsychiatry genes discovered by independent approaches.

### Associations of OOA-enriched protein-coding variants with mood disorders

Association testing in founder populations has the potential to identify population-enriched, functional alleles with substantial effects on disease risk. We therefore annotated the SNPs at each locus to identify protein-coding variants in strong LD with our lead SNPs (D’ > 0.9). For this purpose, we utilized our WGS from the ACP sample so as to include unimputed, population-specific variants. This analysis revealed 15 non-synonymous variants (Table S4). Of these, three stood out based on their strength of linkage with lead SNPs, their enrichment in the OOA compared to the broader European population, and their predicted deleteriousness: chr7:102278021:A:C (rs118010189, CUX1 K500Q), chr16:58576526:C:T (rs201250006, CNOT1 M547I), and chr16:58551644:G:A (rs960417287, CNOT1 A1049V). The latter two variants are in perfect LD in our sample.

*CUX1* encodes Cut Like Homeobox 1, and the rs118010189 variant is located in an alternatively spliced exon that is included only in the CUX1 Alternative Splicing Product (*CASP*) isoform. Previously, rare, protein-truncating and regulatory variants in *CUX1* have been implicated in neurodevelopmental disorders and autism (75). The protein product of the canonical *CUX1* transcript is a homeodomain transcription factor with well-established roles in the development of cortical projection neurons and cerebellar granule neurons (75–78). The *CASP* isoform lacks a DNA-binding domain, interacts biochemically with other CUX1 isoforms (79), and has independent functions as a transmembrane protein involved in intra-Golgi retrograde transport (79–82). Notably, *CUX1* is a hub gene of the OOA mood disorder risk gene network (Fig. 4B). We examined the expression of exons specific to the CUX1 and CASP isoforms in the developing and adult brain using RNA sequencing data from BrainSpan (67) (Fig. S5A). As expected, *CUX1* exons were expressed most highly in the primary visual cortex and in the cerebellum. Intriguingly, *CASP* exons were highly expressed only in the cerebellum and did not have substantial expression in the cortex. These results suggest differential use of *CUX1* and *CASP* isoforms, and that the rs118010189 variant may have its greatest impact in the cerebellum.

*CNOT1* encodes CCR4-NOT Transcription Complex Subunit 1, a component of a transcription factor complex implicated in brain development (83). Pediatric carriers of loss-of-function variants in *CNOT1* have been described to have developmental delay, as well as mental health conditions such as attention deficit and hyperactivity disorder and autism spectrum disorder. One of the few adult carriers in the published case series had bipolar disorder (83). Data from BrainSpan suggest that *CNOT1* is broadly expressed in the brain, with the highest expression at prenatal timepoints (Fig. S5B).

## DISCUSSION

Our findings build on >40 years of research on mood disorders in the OOA population, which previously provided insight into the genetic architecture of mood disorders but were underpowered to detect specific risk loci (22,24–27). Here, in an expanded sample, we identified the first genome-wide significant risk loci for mood disorders in this population. These OOA-specific risk alleles have larger effects than common variants identified in the broader population, most likely explained by founder effects. They act additively with previously described common risk variants for mood disorders and influence sub-clinical behavioral and cognitive traits. Gene network analyses suggest that the loci harbor novel risk genes within gene networks that are shared with neuropsychiatry-related genes identified in the broader population. Annotation of the risk loci revealed missense variants impacting neurodevelopmental genes.

The discovery of OOA-specific risk loci for mood disorders was facilitated by their large effect sizes. Indeed, the major rationale for studies in founder populations is to identify alleles with larger effects than those that can be discovered through GWAS in the broader population. We cannot rule out winner’s curse effects, which would predict that the true effect sizes are smaller than is observed in the current sample. And the lack of an independent cohort is an important limitation. However, large effects are plausible, especially since we find evidence for founder effects. Other factors that may contribute to large effect sizes in the OOA include the relative uniformity in education, lifestyle, and socioeconomic status, and the reduced influence of alcohol and illicit drugs – all of which may provide higher fidelity of neuropsychiatric phenotyping.

Risk for mood disorders in the OOA appears to be highly polygenic, despite the relatively large effects of certain risk loci. Modeling polygenic risk from common variants together with population-specific risk alleles suggested additive contributions. The extent of polygenicity may vary among OOA individuals. It is plausible that the rare 3q28/29 and 5q13 risk alleles confer risk for mood disorders in a pseudo-Mendelian fashion. For carriers of the 7q22 and 16q21 risk alleles, polygenic background and non-genetic factors likely play larger roles. Many OOA individuals with mood disorders are not carriers of any of these population-specific risk alleles. These observations extend previous work in the broader population showing additive effects of polygenic risk scores and copy number variants (84).

Deep phenotyping provided insights into the effects of risk variants on sub-clinical phenotypes. We found that the 3q28/29, 7q22, and 16q21 risk alleles were associated with elevated scores on the Maryland Depression Trait Scale, while the 5q13 risk allele was associated with deficits in digit symbol coding. These associations buttress the primary association of these SNPs with mood disorders. We interpret the stronger effects of these SNPs on MDTS vs. the Beck Depression Inventory to indicate that they more strongly influence lifetime history than current symptoms. The digit symbol coding task primarily measures deficits in information processing speed. This task and other measures of processing speed are among the cognitive tasks most consistently found to be impaired in individuals with bipolar disorder and major depression (85–88). We note that these results are limited by the relatively small sample, which precluded analyses stratified by affection status (i.e., to test whether the SNPs influence these quantitative phenotypes in individuals whose symptoms do not qualify for a major mood disorder). Nonetheless, these findings demonstrate promise for utilizing population isolates to gain insight into the genetic influences on endophenotypes and should be followed up with larger sample sizes and additional sub-clinical assessments.

Network analysis suggested that genes proximal to OOA-specific risk loci converge on a highly polygenic gene network shared with neuropsychiatry risk genes identified by independent approaches. This analysis provided the strongest evidence for interactions with genes that are targets of *CELF4* and *RBFOX1/2/3*, which are neuron-specific RNA binding proteins with roles in neurodevelopment. For instance, *RBFOX1*, encodes RNA Binding Fox-1 Homolog 1, a neuron-specific splicing factor. *RBFOX1* itself is located at a top GWAS risk locus for major depression (6) and is disrupted by copy number variants associated with risk for autism spectrum disorder(89), and its targets have previously been implicated in risk for major depression(6), bipolar disorder (90), schizophrenia (62), and autism (91) through GWAS and exome sequencing studies. These network enrichments support the biological relevance of OOA-specific risk loci -- including those at suggestive levels of significance in our GWAS -- and will aid in the prioritization of specific genes for follow-up studies.

Variant annotation identified promising protein-coding variants at the 7q22 and 16q21 risk loci, in the genes *CUX1* and *CNOT1*. Both these genes are highly plausible candidates with established roles in brain development and previously implicated in risk for neurodevelopmental disorders. However, it is important to note that these variants, if causal, are unlikely to be the only causal variants at these loci. Both loci include hundreds of additional variants, some of which could have important gene regulatory functions, which remain difficult to predict bioinformatically. It is also possible that the risk loci tag structural variants that were not considered in our analysis. The relatively modest enrichment of CUX1 K500Q in the OOA (it has a minor allele frequency of 0.019 in the Amish and 0.007 in the broader European population) puts an upper bound on its true effect size. The *CNOT1* variants are >170-fold enriched in the OOA, with minor allele frequencies less than 0.0001 in the broader population. However, the linkage structure at the 16q21 locus suggests that multiple haplotypes contribute to the signal in this region, with the *CNOT1* variants being much less common than the lead SNP. Nonetheless, these variants represent some of the stronger leads to have emerged from studies of rare variants in mood disorders.

The discovery of OOA-specific risk loci for mood disorders enables a variety of future studies. Additional deep phenotyping may include assessments of brain structure and function. Functional studies may be merited, particularly for *CUX1* and *CNOT1*. Additional loci are likely to be discovered by continuing to expand our sample and through analyses of family-specific variants that could be identified through genome sequencing. These and other family-based studies will continue to provide insights into the etiology and pathophysiological mechanisms of psychiatric disorders, complementary with large-scale GWAS and sequencing studies in the broader population. Family studies are likely the most efficient strategy to characterize specific variants with moderate to large effects.

## ACKNOWLEDGEMENTS

This study was supported by grants and contracts from the National Institute of Mental Health (U01 MH108148 to L.E.H. and P.K., R01 MH110437 to P.P.Z., U01 MH105630 to D.C.G., U01 MH105632 to J.B.), the Regeneron Genetics Center, the Intramural Research Program of the National Institute of Mental Health (ZIA MH002843 to F.J.M.), and a NARSAD Young Investigator Award from the Brain and Behavior Research Foundation to S.A.A. Most of all, we thank the Amish and Mennonite participants, without whose longstanding partnership this study would not have been possible.

## REFERENCES

1. GBD 2017 Disease and Injury Incidence and Prevalence Collaborators (2018): Global, regional, and national incidence, prevalence, and years lived with disability for 354 diseases and injuries for 195 countries and territories, 1990-2017: a systematic analysis for the Global Burden of Disease Study 2017. Lancet Lond Engl 392: 1789–1858.

2. Smoller JW, Finn CT (2003): Family, twin, and adoption studies of bipolar disorder. Am J Med Genet C Semin Med Genet 123C: 48–58.

3. Barnett JH, Smoller JW (2009): The genetics of bipolar disorder. Neuroscience 164: 331–343.

4. Sullivan PF, Neale MC, Kendler KS (2000): Genetic epidemiology of major depression: review and meta-analysis. Am J Psychiatry 157: 1552–1562.

5. Kendler KS, Gatz M, Gardner CO, Pedersen NL (2006): A Swedish national twin study of lifetime major depression. Am J Psychiatry 163: 109–114.

6. Wray NR, Ripke S, Mattheisen M, Trzaskowski M, Byrne EM, Abdellaoui A, et al. (2018): Genome-wide association analyses identify 44 risk variants and refine the genetic architecture of major depression. Nat Genet 50: 668–681.

7. Howard DM, Adams MJ, Clarke T-K, Hafferty JD, Gibson J, Shirali M, et al. (2019): Genome-wide meta-analysis of depression identifies 102 independent variants and highlights the importance of the prefrontal brain regions. Nat Neurosci 22: 343–352.

8. Stahl EA, Breen G, Forstner AJ, McQuillin A, Ripke S, Trubetskoy V, et al. (2019): Genome-wide association study identifies 30 loci associated with bipolar disorder. Nat Genet 51: 793–803.

9. Mullins N, Forstner AJ, O’Connell KS, Coombes B, Coleman JRI, Qiao Z, et al. (2021): Genome-wide association study of more than 40,000 bipolar disorder cases provides new insights into the underlying biology. Nat Genet 53: 817–829.

10. Levey DF, Stein MB, Wendt FR, Pathak GA, Zhou H, Aslan M, et al. (2021): Bi-ancestral depression GWAS in the Million Veteran Program and meta-analysis in >1.2 million individuals highlight new therapeutic directions. Nat Neurosci 24: 954–963.

11. Cross-Disorder Group of the Psychiatric Genomics Consortium. Electronic address, Cross-Disorder Group of the Psychiatric Genomics Consortium (2019): Genomic Relationships, Novel Loci, and Pleiotropic Mechanisms across Eight Psychiatric Disorders. Cell 179: 1469-1482.e11.

12. Cross-Disorder Group of the Psychiatric Genomics Consortium, Lee SH, Ripke S, Neale BM, Faraone SV, Purcell SM, et al. (2013): Genetic relationship between five psychiatric disorders estimated from genome-wide SNPs. Nat Genet 45: 984–994.

13. Ament SA, Szelinger S, Glusman G, Ashworth J, Hou L, Akula N, et al. (2015): Rare variants in neuronal excitability genes influence risk for bipolar disorder. Proc Natl Acad Sci U S A 112: 3576–3581.

14. Strauss KA, Puffenberger EG (2009): Genetics, medicine, and the Plain people. Annu Rev Genomics Hum Genet 10: 513–536.

15. Hou L, Faraci G, Chen DTW, Kassem L, Schulze TG, Shugart YY, McMahon FJ (2013): Amish revisited: next-generation sequencing studies of psychiatric disorders among the Plain people. Trends Genet TIG 29: 412–418.

16. Lopes FL, Hou L, Boldt ABW, Kassem L, Alves VM, Nardi AE, McMahon FJ (2016): Finding Rare, Disease-Associated Variants in Isolated Groups: Potential Advantages of Mennonite Populations. Hum Biol 88: 109–120.

17. Hostetler JA (1993): Amish Society, 4th ed. Baltimore: Johns Hopkins University Press.

18. Smith C (2012): The Mennonites: A Brief History of Their Origins and Later Development in Both Europe and America. Hard Press Publishing.

19. Krahn C, Bender H, Friesen J (1989): Migrations. Global Anabaptist Mennonite Encyclopedia Online. Retrieved November 13, 2020, from https://gameo.org/index.php?title=Migrations&oldid=143668

20. Mckusick VA, Hostetler JA, Egeland JA (1964): GENETIC STUDIES OF THE AMISH, BACKGROUND AND POTENTIALITIES. Bull Johns Hopkins Hosp 115: 203–222.

21. Strauss KA, Markx S, Georgi B, Paul SM, Jinks RN, Hoshi T, et al. (2014): A population-based study of KCNH7 p.Arg394His and bipolar spectrum disorder. Hum Mol Genet 23: 6395–6406.

22. Georgi B, Craig D, Kember RL, Liu W, Lindquist I, Nasser S, et al. (2014): Genomic view of bipolar disorder revealed by whole genome sequencing in a genetic isolate. PLoS Genet 10: e1004229.

23. Pollin TI, Damcott CM, Shen H, Ott SH, Shelton J, Horenstein RB, et al. (2008): A null mutation in human APOC3 confers a favorable plasma lipid profile and apparent cardioprotection. Science 322: 1702–1705.

24. Egeland JA, Gerhard DS, Pauls DL, Sussex JN, Kidd KK, Allen CR, et al. (1987): Bipolar affective disorders linked to DNA markers on chromosome 11. Nature 325: 783–787.

25. Kelsoe JR, Ginns EI, Egeland JA, Gerhard DS, Goldstein AM, Bale SJ, et al. (1989): Re-evaluation of the linkage relationship between chromosome 11p loci and the gene for bipolar affective disorder in the Old Order Amish. Nature 342: 238–243.

26. Kember RL, Hou L, Ji X, Andersen LH, Ghorai A, Estrella LN, et al. (2018): Genetic pleiotropy between mood disorders, metabolic, and endocrine traits in a multigenerational pedigree. Transl Psychiatry 8: 218.

27. Kember RL, Georgi B, Bailey-Wilson JE, Stambolian D, Paul SM, Bućan M (2015): Copy number variants encompassing Mendelian disease genes in a large multigenerational family segregating bipolar disorder. BMC Genet 16: 27.

28. Kessler MD, Loesch DP, Perry JA, Heard-Costa NL, Taliun D, Cade BE, et al. (2020): De novo mutations across 1,465 diverse genomes reveal mutational insights and reductions in the Amish founder population. Proc Natl Acad Sci U S A 117: 2560–2569.

29. Taliun D, Harris DN, Kessler MD, Carlson J, Szpiech ZA, Torres R, et al. (2021): Sequencing of 53,831 diverse genomes from the NHLBI TOPMed Program. Nature 590: 290–299.

30. Purcell SM, Chang CC (n.d.): PLINK v1.9. Retrieved from www.cog-genomics.org/plink/1.9

31. Chang CC, Chow CC, Tellier LC, Vattikuti S, Purcell SM, Lee JJ (2015): Second-generation PLINK: rising to the challenge of larger and richer datasets. GigaScience 4: 7.

32. (N.d.): Retrieved from https://imputation.sanger.ac.uk/

33. Das S, Forer L, Schönherr S, Sidore C, Locke AE, Kwong A, et al. (2016): Next-generation genotype imputation service and methods. Nat Genet 48: 1284–1287.

34. Li H (2011): A statistical framework for SNP calling, mutation discovery, association mapping and population genetical parameter estimation from sequencing data. Bioinforma Oxf Engl 27: 2987–2993.

35. Garrison E, Marth G (2012): Haplotype-based variant detection from short-read sequencing. ArXiv12073907 Q-Bio. Retrieved December 21, 2021, from http://arxiv.org/abs/1207.3907

36. Fernandez-Pujals AM, Adams MJ, Thomson P, McKechanie AG, Blackwood DHR, Smith BH, et al. (2015): Epidemiology and Heritability of Major Depressive Disorder, Stratified by Age of Onset, Sex, and Illness Course in Generation Scotland: Scottish Family Health Study (GS:SFHS). PloS One 10: e0142197.

37. Lichtenstein P, Yip BH, Björk C, Pawitan Y, Cannon TD, Sullivan PF, Hultman CM (2009): Common genetic determinants of schizophrenia and bipolar disorder in Swedish families: a population-based study. Lancet Lond Engl 373: 234–239.

38. Hulshoff Pol HE, van Baal GCM, Schnack HG, Brans RGH, van der Schot AC, Brouwer RM, et al. (2012): Overlapping and segregating structural brain abnormalities in twins with schizophrenia or bipolar disorder. Arch Gen Psychiatry 69: 349–359.

39. Kang HM, Sul JH, Service SK, Zaitlen NA, Kong S-Y, Freimer NB, et al. (2010): Variance component model to account for sample structure in genome-wide association studies. Nat Genet 42: 348–354.

40. Hellevik O (2009): Linear versus logistic regression when the dependent variable is a dichotomy. Qual Quant 43: 59–74.

41. Quinlan AR (2014): BEDTools: The Swiss-Army Tool for Genome Feature Analysis. Curr Protoc Bioinforma 47: 11.12.1-34.

42. Schizophrenia Working Group of the Psychiatric Genomics Consortium (2014): Biological insights from 108 schizophrenia-associated genetic loci. Nature 511: 421–427.

43. Lee JJ, Wedow R, Okbay A, Kong E, Maghzian O, Zacher M, et al. (2018): Gene discovery and polygenic prediction from a genome-wide association study of educational attainment in 1.1 million individuals. Nat Genet 50: 1112–1121.

44. Gandal MJ, Zhang P, Hadjimichael E, Walker RL, Chen C, Liu S, et al. (2018): Transcriptome-wide isoform-level dysregulation in ASD, schizophrenia, and bipolar disorder. Science 362: eaat8127.

45. Singh T, Neale BM, Daly MJ (2020): Exome Sequencing Identifies Rare Coding Variants in 10 Genes Which Confer Substantial Risk for Schizophrenia. Genetic and Genomic Medicine. https://doi.org/10.1101/2020.09.18.20192815

46. Satterstrom FK, Kosmicki JA, Wang J, Breen MS, De Rubeis S, An J-Y, et al. (2020): Large-Scale Exome Sequencing Study Implicates Both Developmental and Functional Changes in the Neurobiology of Autism. Cell 180: 568-584.e23.

47. Abrahams BS, Arking DE, Campbell DB, Mefford HC, Morrow EM, Weiss LA, et al. (2013): SFARI Gene 2.0: a community-driven knowledgebase for the autism spectrum disorders (ASDs). Mol Autism 4: 36.

48. Choi SW, O’Reilly PF (2019): PRSice-2: Polygenic Risk Score software for biobank-scale data. GigaScience 8: giz082.

49. R Core Team (2019): R: A Language and Environment for Statistical Computing. Vienna, Austria: R Foundation for Statistical Computing.

50. Beck AT, Steer RA, Brown GK (1996): BDI-II, Beck Depression Inventory: Manual, 2nd ed. San Antonio, Tex.□: Boston: Psychological Corp.□; Harcourt Brace.

51. Chiappelli J, Nugent KL, Thangavelu K, Searcy K, Hong LE (2014): Assessment of trait and state aspects of depression in schizophrenia. Schizophr Bull 40: 132–142.

52. Bruce HA, Kochunov P, Mitchell B, Strauss KA, Ament SA, Rowland LM, et al. (2019): Clinical and genetic validity of quantitative bipolarity. Transl Psychiatry 9: 228.

53. Wechsler D (2011): Wechsler Abbreviated Scale of Intelligence: WASI-II□; Manual, 2. ed. Bloomington, Minn: Pearson.

54. Ganjgahi H, Winkler AM, Glahn DC, Blangero J, Kochunov P, Nichols TE (2015): Fast and powerful heritability inference for family-based neuroimaging studies. NeuroImage 115: 256–268.

55. de Leeuw CA, Mooij JM, Heskes T, Posthuma D (2015): MAGMA: generalized gene-set analysis of GWAS data. PLoS Comput Biol 11: e1004219.

56. Hasin N, Riggs LM, Shekhtman T, Ashworth J, Lease R, Oshone RT, et al. (2021): A Rare Variant in D-Amino Acid Oxidase Implicates NMDA Receptor Signaling and Cerebellar Gene Networks in Risk for Bipolar Disorder. Psychiatry and Clinical Psychology. https://doi.org/10.1101/2021.06.02.21258261

57. Casella AM, Colantuoni C, Ament SA (2021): Regulome-Wide Association Study Identifies Enhancer Properties Associated with Risk for Schizophrenia. Genomics. https://doi.org/10.1101/2021.06.14.448418

58. Wang D, Liu S, Warrell J, Won H, Shi X, Navarro FCP, et al. (2018): Comprehensive functional genomic resource and integrative model for the human brain. Science 362: eaat8464.

59. Luciano M, Hagenaars SP, Davies G, Hill WD, Clarke T-K, Shirali M, et al. (2018): Association analysis in over 329,000 individuals identifies 116 independent variants influencing neuroticism. Nat Genet 50: 6–11.

60. Wright CF, Fitzgerald TW, Jones WD, Clayton S, McRae JF, van Kogelenberg M, et al. (2015): Genetic diagnosis of developmental disorders in the DDD study: a scalable analysis of genome-wide research data. Lancet Lond Engl 385: 1305–1314.

61. Karczewski KJ, Francioli LC, Tiao G, Cummings BB, Alföldi J, Wang Q, et al. (2020): The mutational constraint spectrum quantified from variation in 141,456 humans. Nature 581: 434–443.

62. Genovese G, Fromer M, Stahl EA, Ruderfer DM, Chambert K, Landén M, et al. (2016): Increased burden of ultra-rare protein-altering variants among 4,877 individuals with schizophrenia. Nat Neurosci 19: 1433–1441.

63. Pirooznia M, Wang T, Avramopoulos D, Valle D, Thomas G, Huganir RL, et al. (2012): SynaptomeDB: an ontology-based knowledgebase for synaptic genes. Bioinforma Oxf Engl 28: 897–899.

64. Szklarczyk D, Gable AL, Lyon D, Junge A, Wyder S, Huerta-Cepas J, et al. (2019): STRING v11: protein-protein association networks with increased coverage, supporting functional discovery in genome-wide experimental datasets. Nucleic Acids Res 47: D607–D613.

65. Csardi G, Nepusz T (2006): The igraph software package for complex network research. Complex Syst.

66. Huang DW, Sherman BT, Lempicki RA (2009): Systematic and integrative analysis of large gene lists using DAVID bioinformatics resources. Nat Protoc 4: 44–57.

67. Li M, Santpere G, Imamura Kawasawa Y, Evgrafov OV, Gulden FO, Pochareddy S, et al. (2018): Integrative functional genomic analysis of human brain development and neuropsychiatric risks. Science 362: eaat7615.

68. Purcell S, Cherny SS, Sham PC (2003): Genetic Power Calculator: design of linkage and association genetic mapping studies of complex traits. Bioinforma Oxf Engl 19: 149–150.

69. Shen H, Damcott CM, Rampersaud E, Pollin TI, Horenstein RB, McArdle PF, et al. (2010): Familial defective apolipoprotein B-100 and increased low-density lipoprotein cholesterol and coronary artery calcification in the old order amish. Arch Intern Med 170: 1850–1855.

70. Clayton-Smith J, Giblin C, Smith RA, Dunn C, Willatt L (2010): Familial 3q29 microdeletion syndrome providing further evidence of involvement of the 3q29 region in bipolar disorder. Clin Dysmorphol 19: 128–132.

71. Mulle JG (2015): The 3q29 deletion confers >40-fold increase in risk for schizophrenia. Mol Psychiatry 20: 1028–1029.

72. Guo X, Ge T, Xia S, Wu H, Colt M, Xie X, et al. (2021): Atp13a5 Marker Reveals Pericytes of the Central Nervous System in Mice. SSRN Electron J. https://doi.org/10.2139/ssrn.3881359

73. Dunn AR, Stout KA, Ozawa M, Lohr KM, Hoffman CA, Bernstein AI, et al. (2017): Synaptic vesicle glycoprotein 2C (SV2C) modulates dopamine release and is disrupted in Parkinson disease. Proc Natl Acad Sci U S A 114: E2253–E2262.

74. Larsen E, Menashe I, Ziats MN, Pereanu W, Packer A, Banerjee-Basu S (2016): A systematic variant annotation approach for ranking genes associated with autism spectrum disorders. Mol Autism 7: 44.

75. Doan RN, Bae B-I, Cubelos B, Chang C, Hossain AA, Al-Saad S, et al. (2016): Mutations in Human Accelerated Regions Disrupt Cognition and Social Behavior. Cell 167: 341-354.e12.

76. Cubelos B, Sebastián-Serrano A, Beccari L, Calcagnotto ME, Cisneros E, Kim S, et al. (2010): Cux1 and Cux2 regulate dendritic branching, spine morphology, and synapses of the upper layer neurons of the cortex. Neuron 66: 523–535.

77. Li N, Zhao C-T, Wang Y, Yuan X-B (2010): The transcription factor Cux1 regulates dendritic morphology of cortical pyramidal neurons. PloS One 5: e10596.

78. Cubelos B, Briz CG, Esteban-Ortega GM, Nieto M (2015): Cux1 and Cux2 selectively target basal and apical dendritic compartments of layer II-III cortical neurons. Dev Neurobiol 75: 163–172.

79. Lievens PM, Tufarelli C, Donady JJ, Stagg A, Neufeld EJ (1997): CASP, a novel, highly conserved alternative-splicing product of the CDP/cut/cux gene, lacks cut-repeat and homeo DNA-binding domains, and interacts with full-length CDP in vitro. Gene 197: 73–81.

80. Ramdzan ZM, Nepveu A (2014): CUX1, a haploinsufficient tumour suppressor gene overexpressed in advanced cancers. Nat Rev Cancer 14: 673–682.

81. Gillingham AK, Pfeifer AC, Munro S (2002): CASP, the alternatively spliced product of the gene encoding the CCAAT-displacement protein transcription factor, is a Golgi membrane protein related to giantin. Mol Biol Cell 13: 3761–3774.

82. Osterrieder A, Sparkes IA, Botchway SW, Ward A, Ketelaar T, de Ruijter N, Hawes C (2017): Stacks off tracks: a role for the golgin AtCASP in plant endoplasmic reticulum-Golgi apparatus tethering. J Exp Bot 68: 3339–3350.

83. Vissers LELM, Kalvakuri S, de Boer E, Geuer S, Oud M, van Outersterp I, et al. (2020): De Novo Variants in CNOT1, a Central Component of the CCR4-NOT Complex Involved in Gene Expression and RNA and Protein Stability, Cause Neurodevelopmental Delay. Am J Hum Genet 107: 164–172.

84. Bergen SE, Ploner A, Howrigan D, CNV Analysis Group and the Schizophrenia Working Group of the Psychiatric Genomics Consortium, O’Donovan MC, Smoller JW, et al. (2019): Joint Contributions of Rare Copy Number Variants and Common SNPs to Risk for Schizophrenia. Am J Psychiatry 176: 29–35.

85. Glahn DC, Almasy L, Barguil M, Hare E, Peralta JM, Kent JW, et al. (2010): Neurocognitive Endophenotypes for Bipolar Disorder Identified in Multiplex Multigenerational Families. Arch Gen Psychiatry 67: 168.

86. Chaves OC, Lombardo LE, Bearden CE, Woolsey MD, Martinez DM, Barrett JA, et al. (2011): Association of clinical symptoms and neurocognitive performance in bipolar disorder: a longitudinal study: Symptoms and cognition in bipolar disorder. Bipolar Disord 13: 118–123.

87. Austin M-P, Ross M, Murray C, O’Caŕroll RE, Ebmeier KP, Goodwin GM (1992): Cognitive function in major depression. J Affect Disord 25: 21–29.

88. Bora E, Harrison BJ, Yücel M, Pantelis C (2013): Cognitive impairment in euthymic major depressive disorder: a meta-analysis. Psychol Med 43: 2017–2026.

89. Sebat J, Lakshmi B, Malhotra D, Troge J, Lese-Martin C, Walsh T, et al. (2007): Strong association of de novo copy number mutations with autism. Science 316: 445–449.

90. Palmer DS, Howrigan DP, Chapman SB, Adolfsson R, Bass N, Blackwood D, et al. (2021): Exome Sequencing in Bipolar Disorder Reveals Shared Risk Gene AKAP11 with Schizophrenia. Genetic and Genomic Medicine. https://doi.org/10.1101/2021.03.09.21252930

91. Lee J-A, Damianov A, Lin C-H, Fontes M, Parikshak NN, Anderson ES, et al. (2016): Cytoplasmic Rbfox1 Regulates the Expression of Synaptic and Autism-Related Genes. Neuron 89: 113–128.

