## Supplementary Figures for "Genome-wide significant risk loci for mood disorders in the Old Order Amish founder population"

**SUPPLEMENTARY TABLES** (as Excel files)

Table S1: Description of cohorts used in this study.

Table S2. Precision and recall for non-reference imputed genotypes based on comparison to whole-genome sequencing in matched samples. Precision and recall are with respect to the 6.6 million SNPs in the imputed dataset and should be interpreted with regard to the accuracy of imputation for these SNPs. However, it should be noted that additional variants (e.g., very rare SNPs and indels) that can be discovered by genome sequencing are not in the reference panel and are ignored in this analysis.

Table S3. Neuropsychiatric diagnoses of Old Order Amish participants in each cohort. BDI=Bipolar I, BDII=Bipolar II, BD:NOS= Bipolar not otherwise specified (category dropped in DSM-V, thus not used in ACP or AMBiGen cohorts), MDD-R=Major depressive disorder-recurring, SCZ= Schizoaffective or Schizophrenia. Unaffected=no mood disorder diagnosis on Axis I or II. Other diagnosis=diagnosed with something other than mood disorder (e.g., anxiety disorder, single episode major depression). Missing=diagnosis not ascertained. All TOPMed samples were presumed unaffected.

Table S4. Summary statistics and functional annotation of SNPs at each genome-wide significant risk locus. Data are shown for lead SNPs and other SNPs in linkage disequilibrium with a lead SNP in WGS from the ACP cohort (D’ > 0.9).

Table S5. Genetic association summary statistics for lead SNPs in pseudoreplication analyses.

Table S6. Genetic association summary statistics for lead SNPs using alternative affection status models.

Table S7. Overlap between OOA mood disorders risk loci and risk loci from GWAS of psychiatric disorders in independent cohorts.

Table S8. Heritability and summary statistics for behavioral and neurocognitive traits.

Table S9. Associations of lead SNPs from mood disorder risk loci with behavioral and cognitive traits.

Table S10. Gene-based p-values and network centrality scores

Table S11. Network-based enrichment of OOA risk genes for interactions with established neuropsychiatry-related gene sets.

Table S12. Gene Ontology terms enriched among the top 250 OOA risk genes prioritized by p-value and network centrality.

**Supplementary Figure 1. Population structure of Amish and Mennonite populations.** Principal component analyses (PCA) of imputed genotypes from the combined samples in ACP, AMBiGen, ASMAD, and TOPMed. **A**. PCA including both OOA and non-OOA Anabaptists (n=2,570). Plot shows the first two PCs, with samples colored by religious group. Amish samples are further subdivided by home state -- Pennsylvania (PA) or non-PA. The vast majority of PA Amish in our sample are Old Order Amish from Lancaster County (OOA). B. PCA calculated using only the Lancaster OOA individuals (n=1,672). Plot of the first two PCs, with samples colored by study and sequencing or genotyping technology.

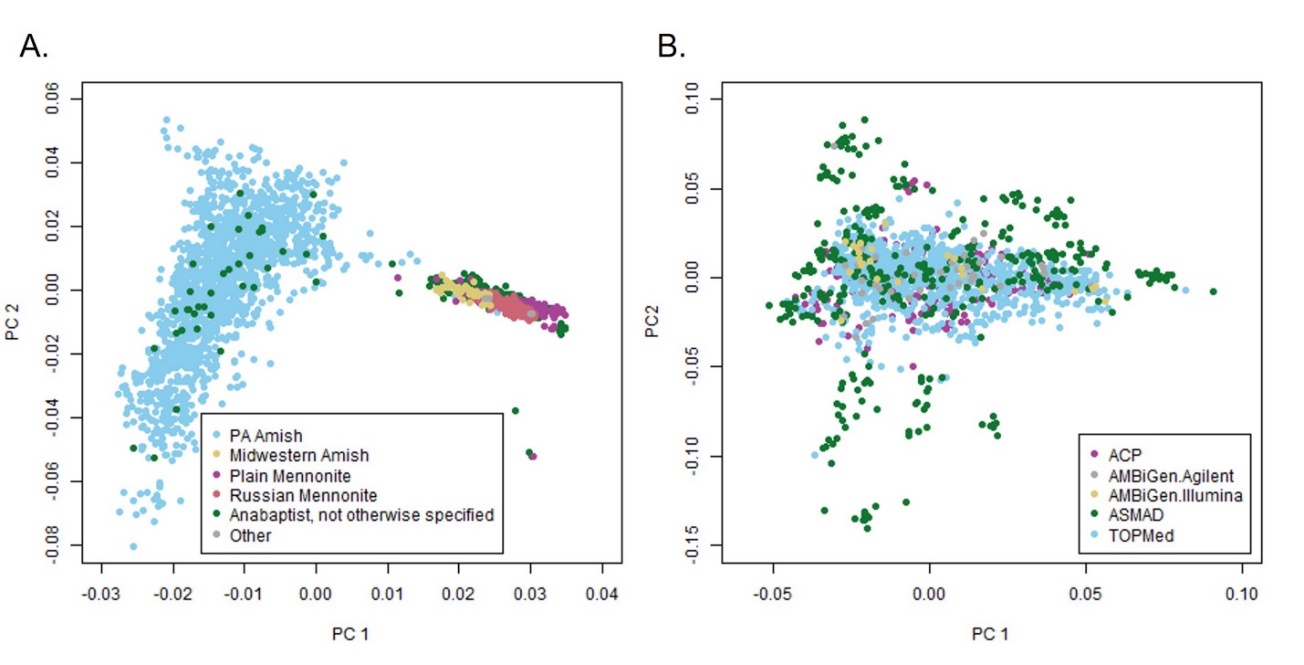

**Supplementary Figure 2. Discovery of risk loci for mood disorders in the Old Order Amish.** A. Quantile-quantile (Q-Q) plot for observed p-values vs. expectation under a null distribution. B. Region plot for locus 3q28/29. C. Region plot for locus 5q13. D. Region plot for locus 7q22. E. Region plot for locus 16q21. Blue dots indicate the lead SNP and SNPs in LD with the lead SNP for each locus. Genomic coordinates of risk loci from published GWAS in the broader population are plotted directly below the x-axis. Positional candidate genes prioritized based on prior evidence from sequencing studies and functional genomics are plotted below each region plot. The orange dot on panel D indicates rs118010189 (CUX1 Lys500Gln).

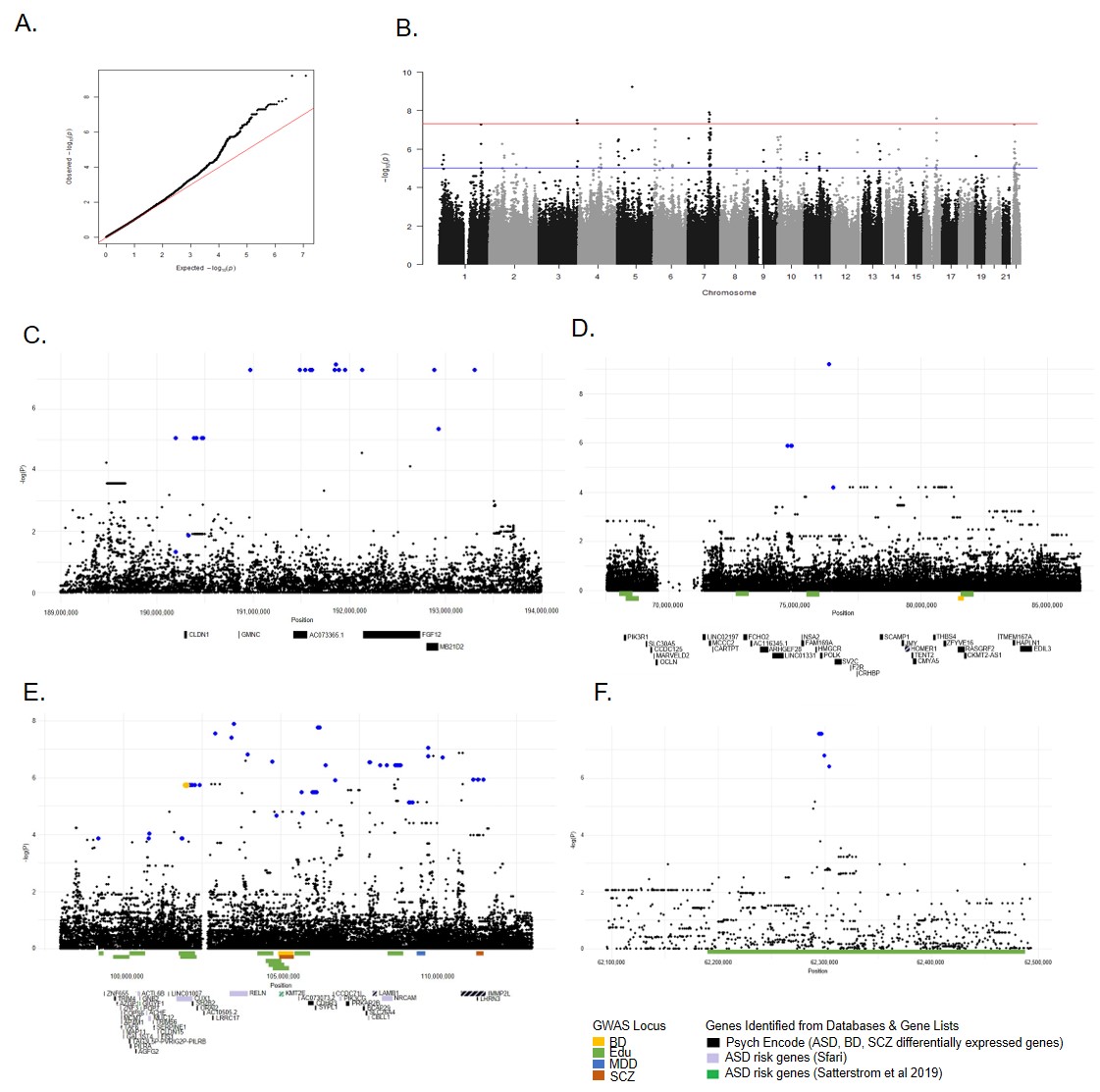

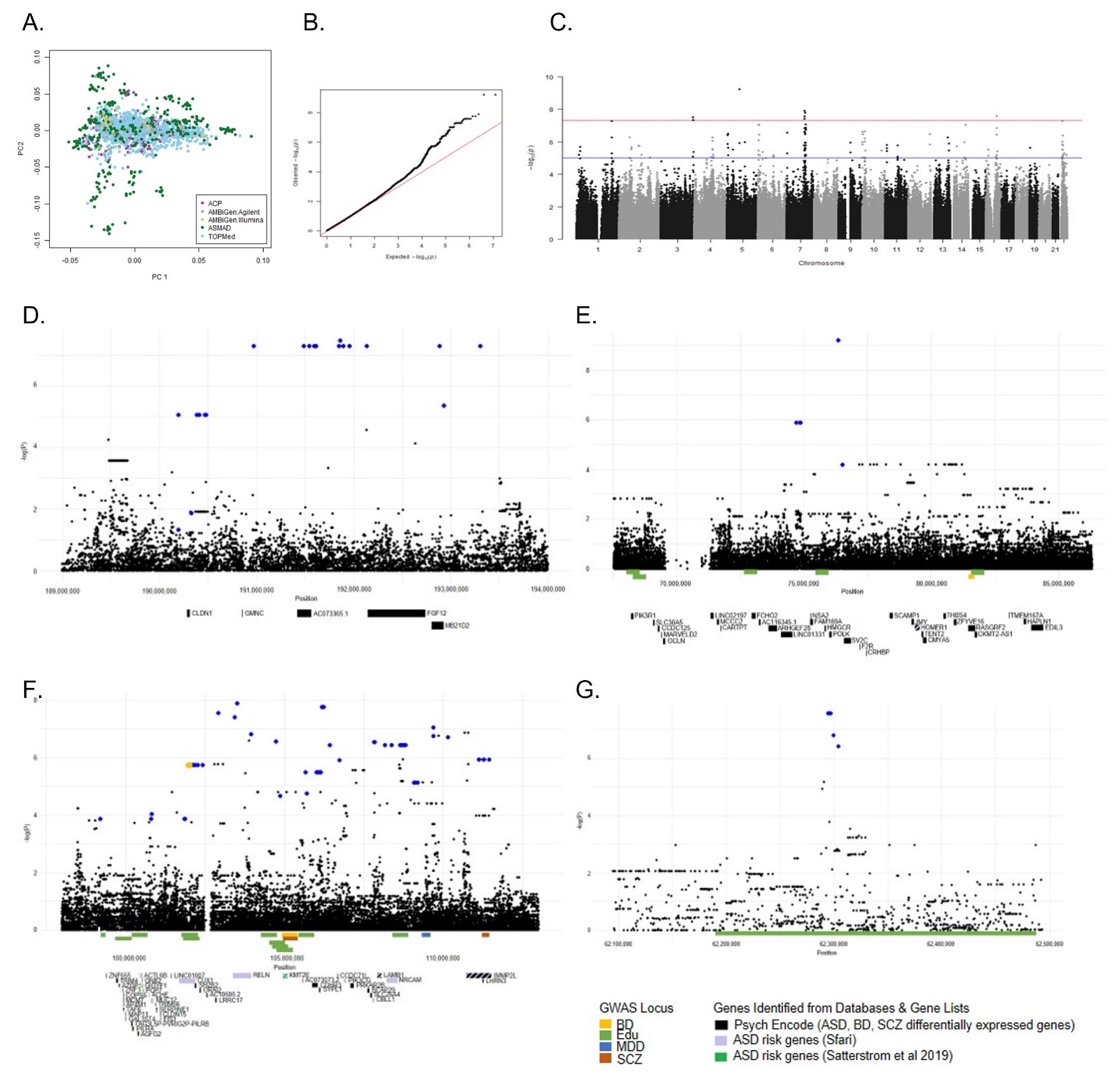

A.

B.

C.

D.

E.

**Supplementary Figure 3. Empirical kinship among carriers of the 3q28/29 and 5q13 risk alleles**. Each plot indicates hierarchical clustering based on an empirical kinship matrix calculated for all 1,672 Lancaster OOA individuals. A. carriers of the 3q28/29 lead SNP. B. carriers of the 5q13 lead SNP. Y-axis indicates the coefficient of relatedness, with the dotted line at 0.5 indicating first-degree relatives. a=ACP, b=AMBiGen, c=ASMAD, d=TOPMed.

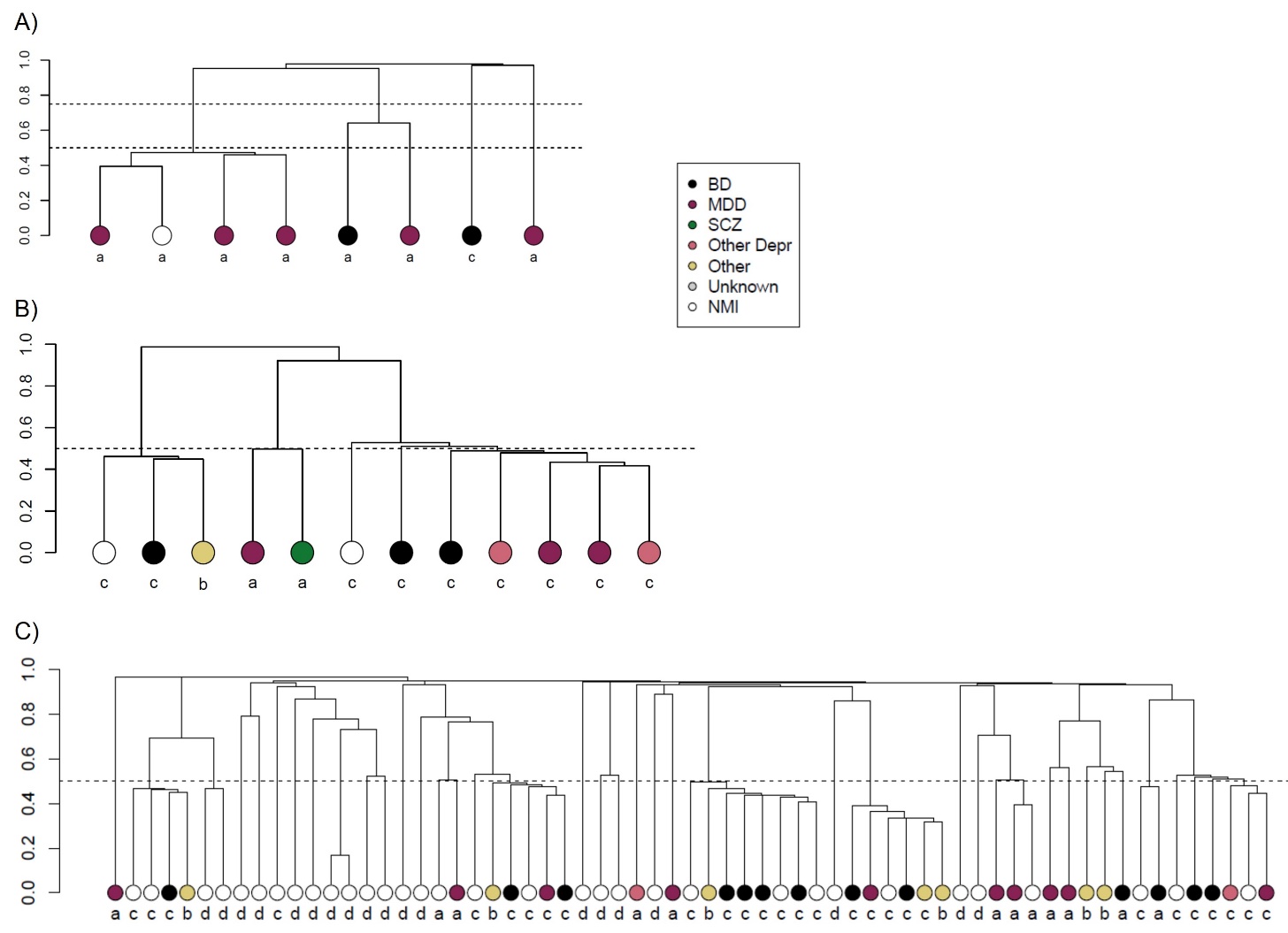

A.

B.

**Supplementary Figure 4. Polygenic risk scores for bipolar disorder, schizophrenia, and major depression.** Polygenic risk scores (PRS) for bipolar disorder, major depressive disorder (MDD) and schizophrenia (SCZ) were derived from published GWAS in the broader European population using PRSice-2 and tested for association with mood disorder affection status in our Old Order Amish sample. A. Disease liability as a function of PRS quartile. B. Distribution of PRS scores in affected individuals, their unaffected relatives, and population controls.

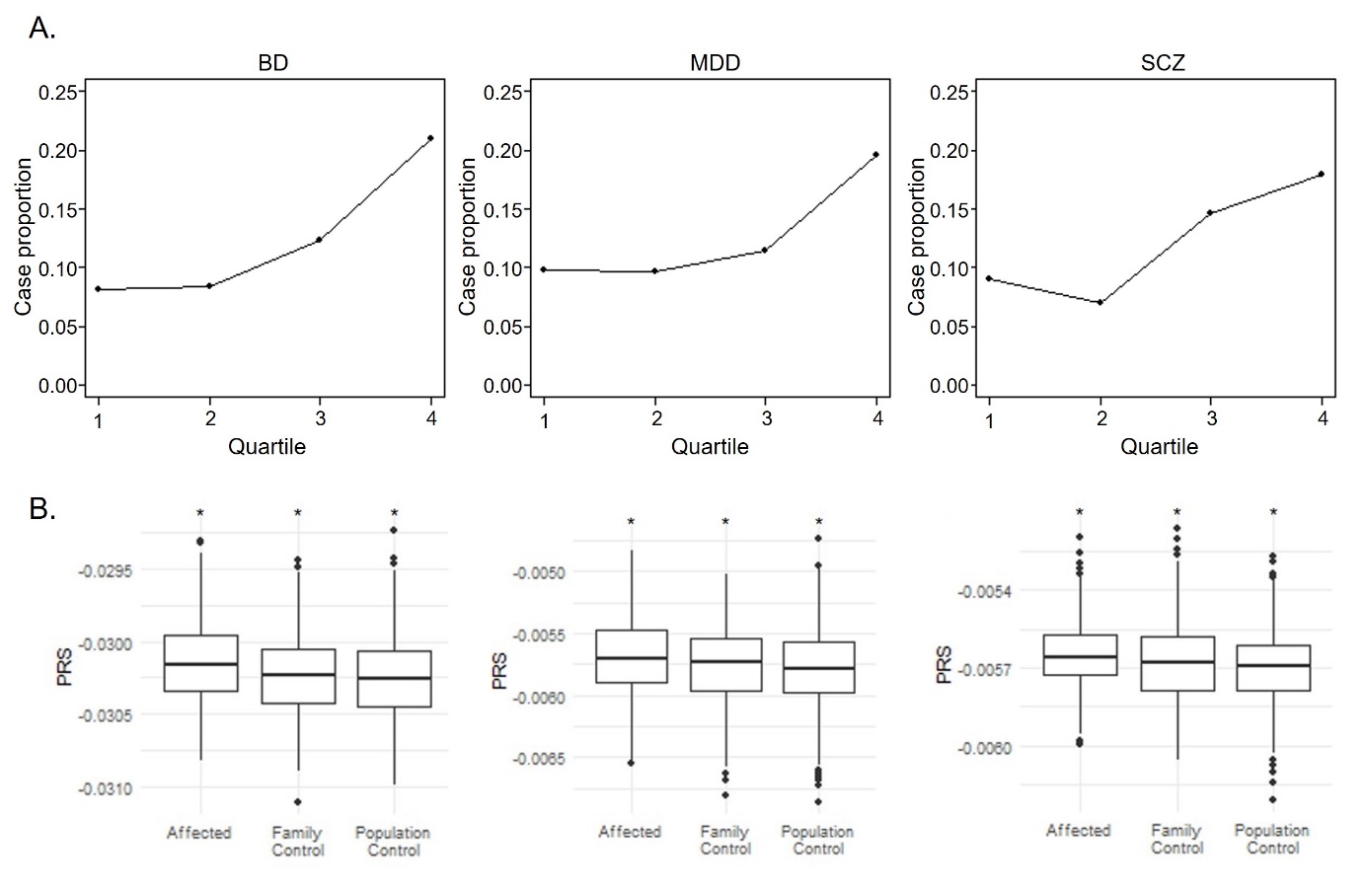

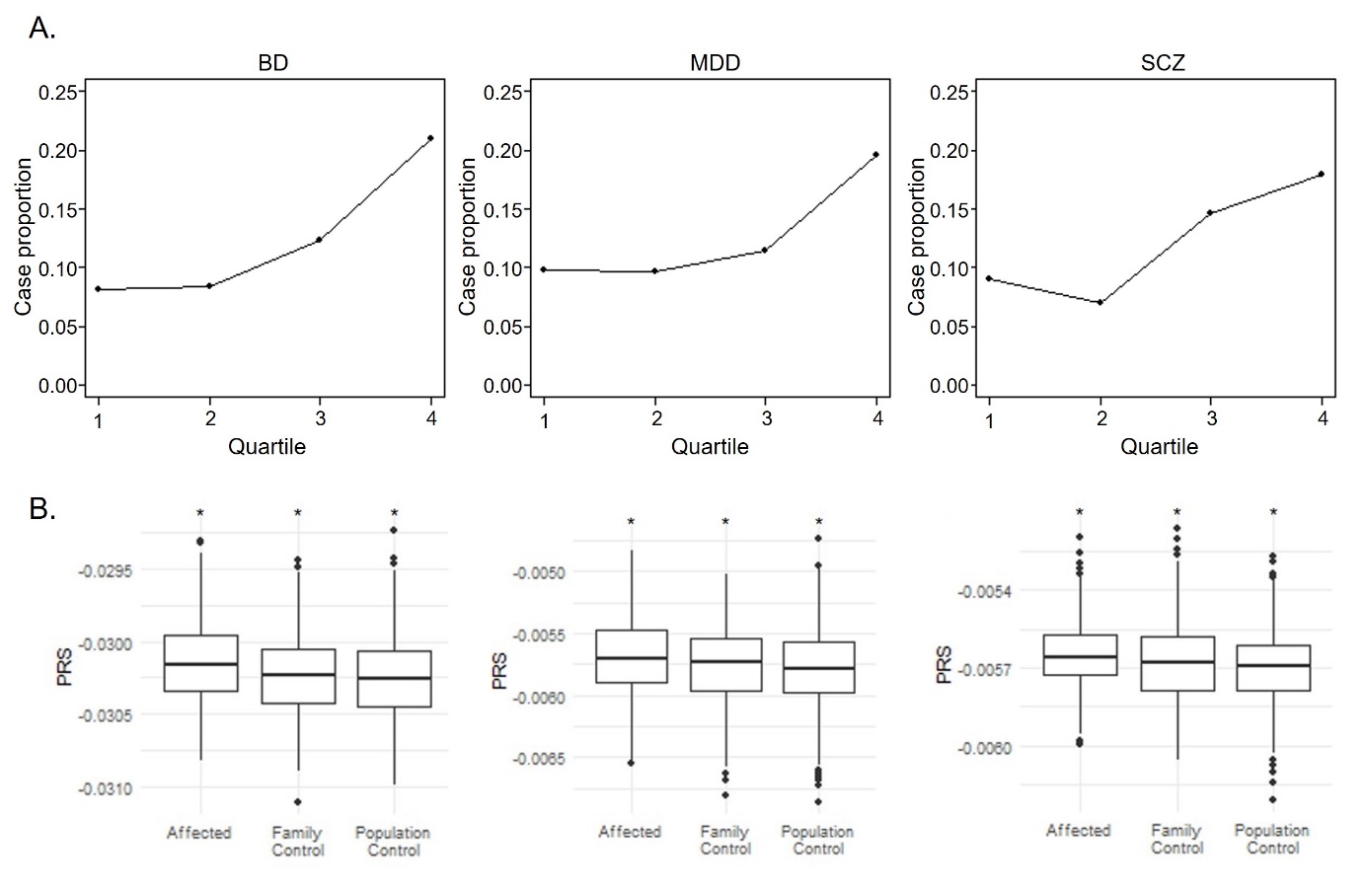

A.

A.

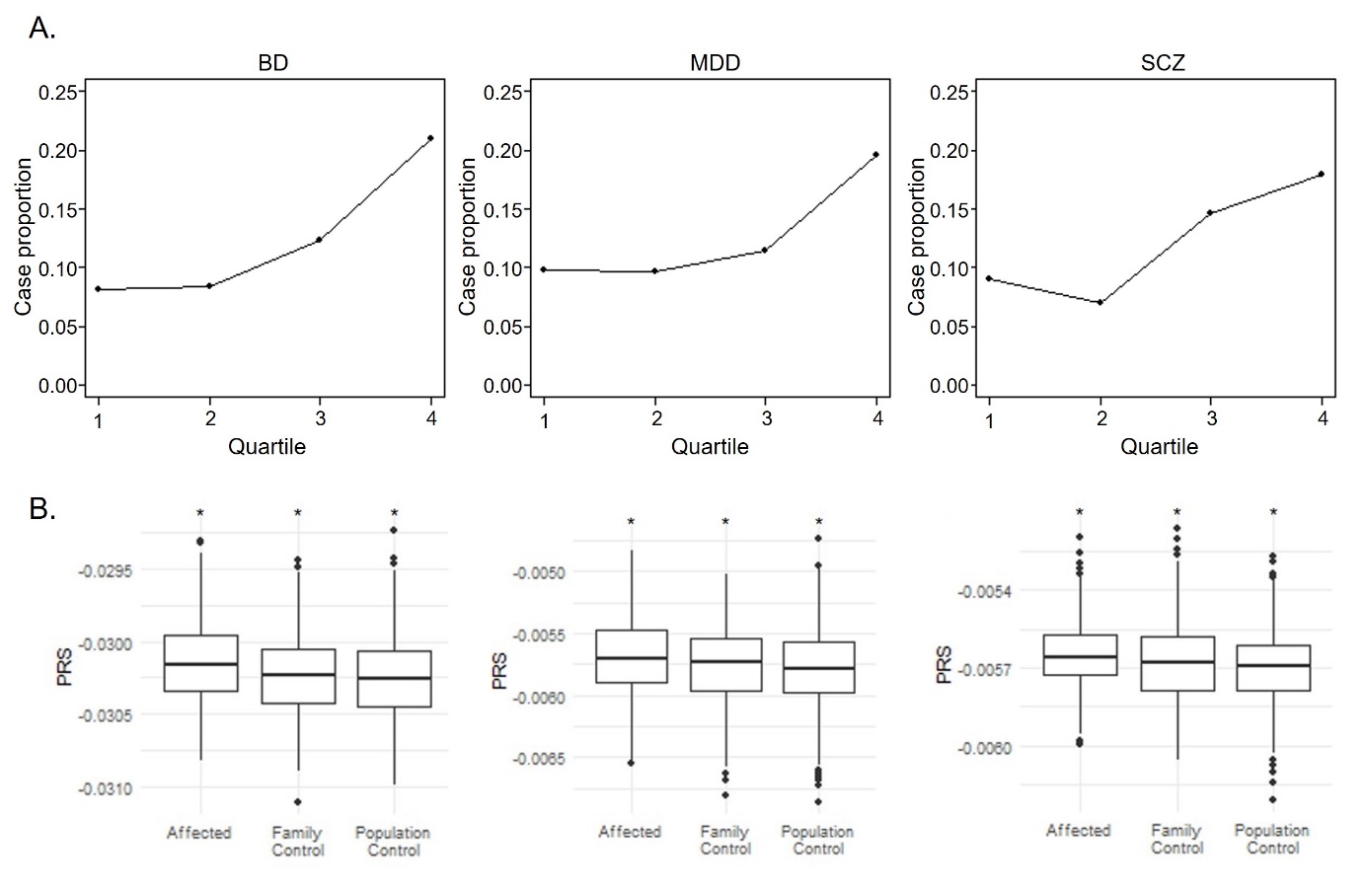

**Supplementary Figure 5. Expression of *CUX1* and *CNOT1* in the human brain.** RNA sequencing data from the BrainSpan Developmental Transcriptome atlas. A. *CUX1/CASP* expression. Data from postnatal samples, summarized to exons. *left*. Expression of a *CUX1*-specific exon across brain regions. *right*. Expression of a *CASP*-specific exon across brain regions. B. *CNOT1* expression. Data summarized to genes. Data shown are for amygdala (AMY), with each point representing a single donor brain. Similar patterns were observed in most brain regions. DFC = dorsolateral prefrontal cortex; VFC = vetrolateral prefrontal cortex; MFC = anterior cingulate (medial prefrontal) cortex; OFC = orbitofrontal cortex; M1C = primary motor cortex; S1C = primary somatosensory cortex; IPC = inferior parietal cortex; A1C = primary auditory cortex; STC = superior temporal cortex; ITC = inferior temporal cortex; V1C = primary visual cortex; HIP = hippocampus; AMY = amygdala; STR = striatum; MD = mediodoral nucleus of the thalamus; CBC = cerebellar cortex.

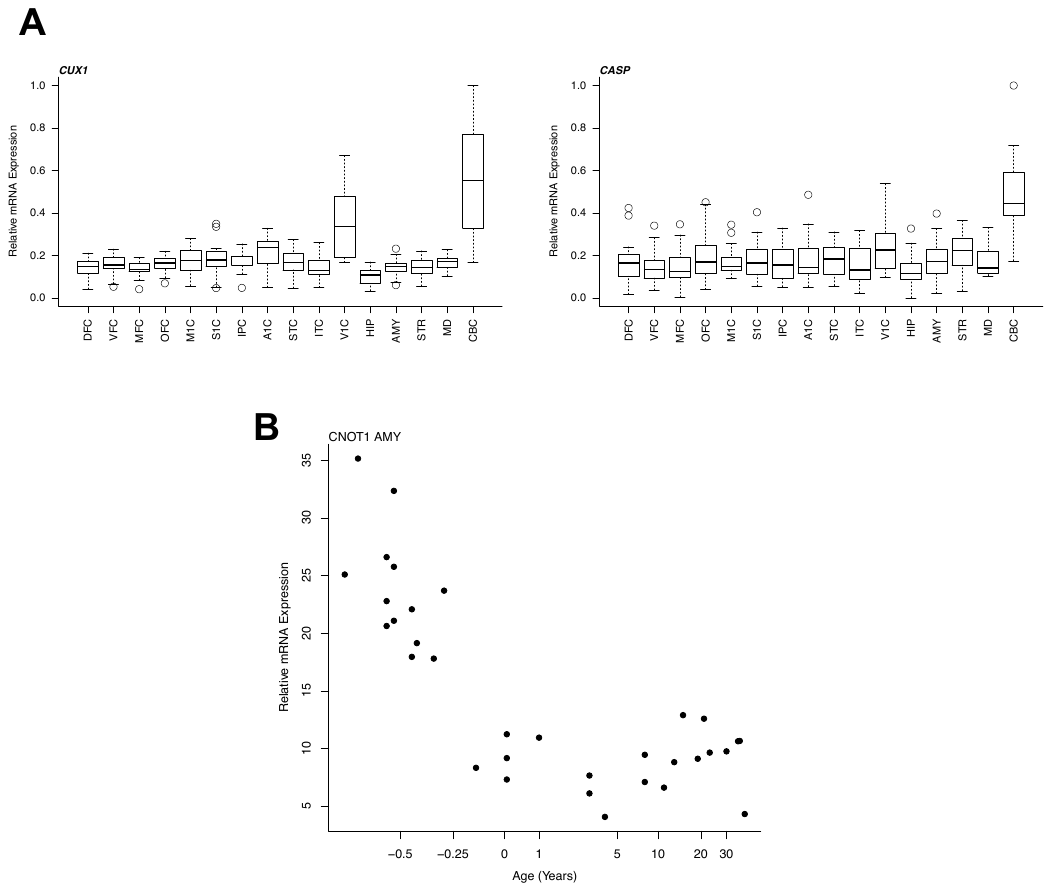
